# Circulating Endothelial Progenitor Cells Reduce the Risk of Alzheimer’s Disease

**DOI:** 10.1101/2023.01.16.23284571

**Authors:** Yixuan Wang, Jinghan Huang, Ting Fang Alvin Ang, Yibo Zhu, Qiushan Tao, Jesse Mez, Michael Alosco, Gerald V. Denis, Anna Belkina, Ashita Gurnani, Mark Ross, Bin Gong, Jingyan Han, Kathryn L. Lunetta, Thor D. Stein, Rhoda Au, Lindsay A. Farrer, Xiaoling Zhang, Wei Qiao Qiu

**Affiliations:** Department of Medicine Biomedical Genetics, Boston University Chobanian & Avedisian School of Medicine, Boston, MA, USA; Department of Medicine Hematology & Medical Oncology, Boston University Chobanian & Avedisian School of Medicine, Boston, MA, USA; Department of Medicine Vascular Biology, Boston University Chobanian & Avedisian School of Medicine, Boston, MA, USA; Department of Medicine Anatomy & Neurobiology, Boston University Chobanian & Avedisian School of Medicine, Boston, MA, USA; Department of Medicine Pharmacology & Experimental Therapeutics, Boston University Chobanian & Avedisian School of Medicine, Boston, MA, USA; Neurology, Boston University Chobanian & Avedisian School of Medicine, Boston, MA, USA; Pathology &Laboratory Medicine, Boston University Chobanian & Avedisian School of Medicine, Boston, MA, USA; Ophthalmology, Boston University Chobanian & Avedisian School of Medicine, Boston, MA, USA; Psychiatry, Boston University Chobanian & Avedisian School of Medicine, Boston, MA, USA; Framingham Heart Study, Boston University Chobanian & Avedisian School of Medicine, Framingham, MA, USA; Alzheimer’s Disease Research Center, Boston University Chobanian & Avedisian School of Medicine, Boston, MA, USA; Department of Biostatistics, Boston University School of Public Health, Boston, MA, USA; Department of Epidemiology, Boston University School of Public Health, Boston, MA, USA; School of Energy, Geosciences, Infrastructure and Society, Institute of Life and Earth Sciences, Heriot-Watt University, Edinburgh, Scotland; VA Boston Healthcare System, Boston, MA, USA; Department of Pathology, University of Texas Medical Branch, Galveston, Texas, USA

**Keywords:** Alzheimer’s disease, CD34+CD133+, circulating endothelial progenitor cells, vascular diseases, GWAS

## Abstract

Cerebrovascular damage coexists with Alzheimer’s disease (AD) pathology and increases AD risk. However, it is unclear whether endothelial progenitor cells reduce AD risk via cerebrovascular repair. By using the Framingham Heart Study (FHS) offspring cohort, which includes data on different progenitor cells, the incidence of AD dementia, peripheral and cerebrovascular pathologies, and genetic data (n = 1,566), we found that elevated numbers of circulating endothelial progenitor cells with CD34+CD133+ co-expressions had a dose-dependent association with decreased AD risk (HR = 0.67, 95% CI: 0.46-0.96, p = 0.03) after adjusting for age, sex, years of education, and *APOE* ε4. With stratification, this relationship was only significant among those individuals who had vascular pathologies, especially hypertension (HTN) and cerebral microbleeds (CMB), but not among those individuals who had neither peripheral nor central vascular pathologies. We applied a genome-wide association study (GWAS) and found that the number of CD34+CD133+ cells impacted AD risk depending on the homozygous genotypes of two genes: *KIRREL3* rs580382 CC carriers (HR = 0.31, 95% CI: 0.17-0.57, p<0.001), *KIRREL3* rs4144611 TT carriers (HR = 0.29, 95% CI: 0.15-0.57, p<0.001), and *EXOC6B* rs61619102 CC carriers (HR = 0.49, 95% CI: 0.31-0.75, p<0.001) after adjusting for confounders. In contrast, the relationship did not exist in their counterpart genotypes, e.g. *KIRREL3* TT/CT or GG/GT carriers and *EXOC6B* GG/GC carriers. Our findings suggest that circulating CD34+CD133+ endothelial progenitor cells can be therapeutic in reducing AD risk in the presence of cerebrovascular pathology, especially in *KIRREL3* and *EXOC6B* genotype carriers.

## Introduction

Consistent with the observation that cerebrovascular pathologies co-exist with neuropathological hallmarks of Alzheimer’s disease (AD), both peripheral and central vascular diseases, including hypertension (HTN) and cerebral microbleeds (CMB), have been shown to increase the risk of late-onset AD.^1^ Microvascular endothelia, which are composed of blood-facing cells of the blood-brain barrier (BBB) in brain capillary beds, may play an important role in the development of AD.^2, 3^ Recently, we demonstrated that monomeric C-reactive protein (mCRP) and apolipoprotein E (*APOE*) protein compete to bind with endothelial membrane surface CD31 protein during peripheral inflammation. ^4^ Although mCRP-CD31 binding causes shortened microvasculature in the brain, thus leading to AD pathology and cognitive impairment, *APOE*-CD31 binding in the presence of the *APOE2* genotype is protective for brain endothelia and against the pathogenesis of AD. Due to the fact that *APOE* ε4 carriers are vulnerable to peripheral inflammation, as a risk factor, leading to endothelial dysfunction, cerebrovascular pathology and AD pathogenesis,^4^ we hypothesized that endothelial progenitor cells, as an antagonizing element, can lessen AD risk in persons with particular AD-associated genotypes.

Blood circulating endothelial progenitor cells have different subtypes and express CD34, CD133, or both proteins. When CD133, a member of a novel family of cell surface glycoproteins, is expressed in hematopoietic-derived CD34+ stem cells, these cells can differentiate into endothelia, vascular smooth muscle cells, and neurons and glia in the brain.^5^ Some studies have suggested that CD34+CD133+ cells are prognostic biomarkers^6^ and can be therapeutic for ischemic diseases.^7–9^ Two studies with a small sample size showed that AD patients had a lower number of circulating CD34+CD133+ cells^10^ or CD34+ cells^11^ than controls. However, it is unknown whether CD34+CD133+ cells are related to AD risk, especially in the presence of cerebrovascular pathology.

In this study, we examined the relationship between different circulating endothelial progenitor cells, endothelial mature cells and other stem cells, vascular pathologies and AD/dementia risk in participants of the Framingham Heart Study (FHS) Offspring Generation 2 (Gen 2) cohort who have been longitudinally examined for AD dementia incidence. It was found that only CD34+CD133+ endothelial progenitor cells reduced AD risk. We further conducted a genome-wide association study (GWAS) to identify loci that are associated with AD risk through their interaction with CD34+CD133+ endothelial progenitor cells. We used the RNAseq expression data from the Religious Orders Study/Memory and Aging Project (ROSMAP) study to explore the relationship between the expressions of the identified genes and AD.

## Results

### Circulating CD34+CD133+ progenitor cells and their associated characteristics in the FHS study

This study included 1,566 participants from the FHS Gen2 cohort with an average age (mean±SD) of 65.93±8.91 years at Exam 8, of which 46.36% were females. The median frequency of circulating CD34+CD133+ progenitor cells was 0.032% mononuclear cells measured at baseline (Exam 8). The participants were then divided into four quartiles based on the circulating CD34+CD133+ frequency (Q1 = 0.002-0.020; Q2 = 0.020-0.032; Q3 = 0.032-0.049 and Q4 = 0.049-0.609). We found that younger (p = 0.007) and female (p = 0.007) participants were more likely to be in the higher CD34+CD133+ frequency quartile (**Table 1**), with no significant differences in education and *APOE* ε4 carriers across the four quartiles. Vascular disease rates also did not significantly differ across the four CD34+CD133+ quartiles.

**Table 1.**
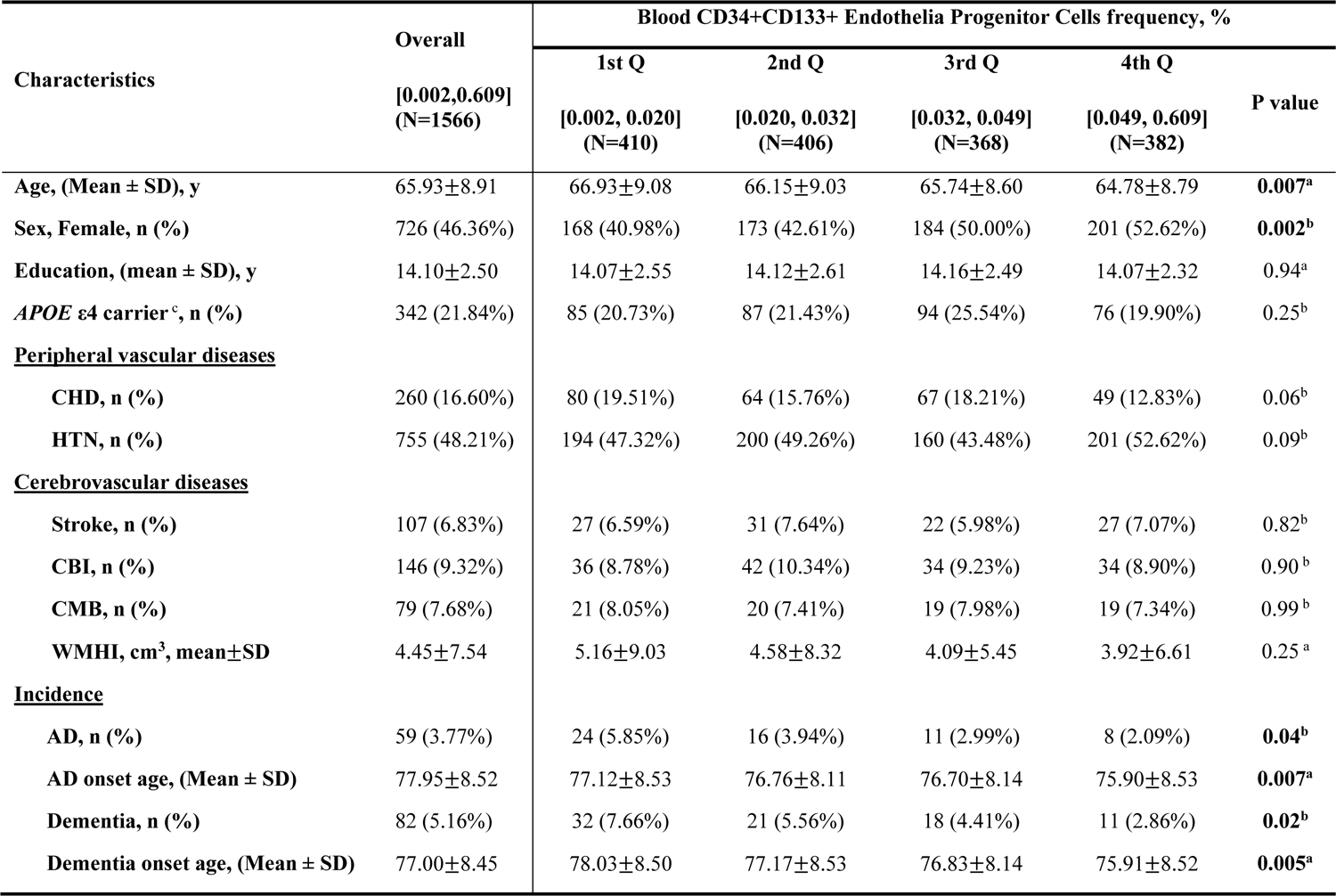
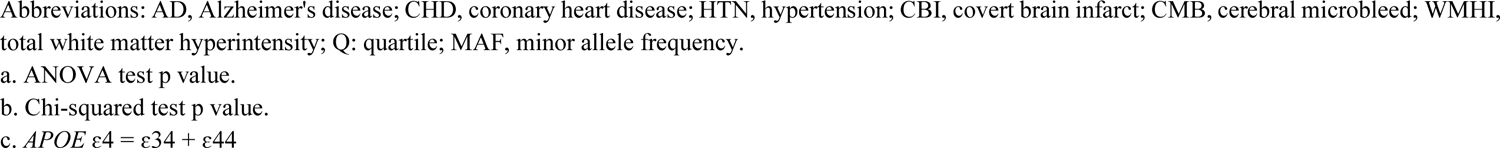
Characteristics of the study sample based on circulating CD34+CD133+ stem cell quartiles

| Characteristics | Overall<br>[0.002,0.609]<br>(N=1566) | Blood CD34+CD133+ Endothelia Progenitor Cells frequency, % |  |  |  | P value |
| --- | --- | --- | --- | --- | --- | --- |
|  |  | 1st Q<br>[0.002, 0.020]<br>(N=410) | 2nd Q<br>[0.020, 0.032]<br>(N=406) | 3rd Q<br>[0.032, 0.049]<br>(N=368) | 4th Q<br>[0.049, 0.609]<br>(N=382) |  |
| Age, (Mean ± SD), y | 65.93±8.91 | 66.93±9.08 | 66.15±9.03 | 65.74±8.60 | 64.78±8.79 | **0.007<sup>a</sup>** |
| Sex, Female, n (%) | 726 (46.36%) | 168 (40.98%) | 173 (42.61%) | 184 (50.00%) | 201 (52.62%) | **0.002<sup>b</sup>** |
| Education, (mean ± SD), y | 14.10±2.50 | 14.07±2.55 | 14.12±2.61 | 14.16±2.49 | 14.07±2.32 | 0.94<sup>a</sup> |
| APOE ε4 carrier <sup>c</sup>, n (%) | 342 (21.84%) | 85 (20.73%) | 87 (21.43%) | 94 (25.54%) | 76 (19.90%) | 0.25<sup>b</sup> |
| **<u>Peripheral vascular diseases</u>** |  |  |  |  |  |  |
| CHD, n (%) | 260 (16.60%) | 80 (19.51%) | 64 (15.76%) | 67 (18.21%) | 49 (12.83%) | 0.06<sup>b</sup> |
| HTN, n (%) | 755 (48.21%) | 194 (47.32%) | 200 (49.26%) | 160 (43.48%) | 201 (52.62%) | 0.09<sup>b</sup> |
| **<u>Cerebrovascular diseases</u>** |  |  |  |  |  |  |
| Stroke, n (%) | 107 (6.83%) | 27 (6.59%) | 31 (7.64%) | 22 (5.98%) | 27 (7.07%) | 0.82<sup>b</sup> |
| CBI, n (%) | 146 (9.32%) | 36 (8.78%) | 42 (10.34%) | 34 (9.23%) | 34 (8.90%) | 0.90<sup>b</sup> |
| CMB, n (%) | 79 (7.68%) | 21 (8.05%) | 20 (7.41%) | 19 (7.98%) | 19 (7.34%) | 0.99<sup>b</sup> |
| WMHI, cm<sup>3</sup>, mean±SD | 4.45±7.54 | 5.16±9.03 | 4.58±8.32 | 4.09±5.45 | 3.92±6.61 | 0.25<sup>a</sup> |
| **<u>Incidence</u>** |  |  |  |  |  |  |
| AD, n (%) | 59 (3.77%) | 24 (5.85%) | 16 (3.94%) | 11 (2.99%) | 8 (2.09%) | **0.04<sup>b</sup>** |
| AD onset age, (Mean ± SD) | 77.95±8.52 | 77.12±8.53 | 76.76±8.11 | 76.70±8.14 | 75.90±8.53 | **0.007<sup>a</sup>** |
| Dementia, n (%) | 82 (5.16%) | 32 (7.66%) | 21 (5.56%) | 18 (4.41%) | 11 (2.86%) | **0.02<sup>b</sup>** |
| Dementia onset age, (Mean ± SD) | 77.00±8.45 | 78.03±8.50 | 77.17±8.53 | 76.83±8.14 | 75.91±8.52 | **0.005<sup>a</sup>** |

Abbreviations: AD, Alzheimer's disease; CHD, coronary heart disease; HTN, hypertension; CBI, covert brain infarct; CMB, cerebral microbleed; WMHI, total white matter hyperintensity; Q: quartile; MAF, minor allele frequency.
a. ANOVA test p value.
b. Chi-squared test p value.
c. *APOE* $\epsilon 4 = \epsilon 34 + \epsilon 44$

### The association between increased CD34+CD133+ endothelial progenitor cells and reduced AD/dementia risk

There were 59 cases of incident AD after exam 8. Interestingly, the incident rates of AD (5.85% vs. 3.94% vs. 2.99% vs. 2.08%, p = 0.036) and all-cause dementia (7.66% vs. 5.56% vs. 4.41% vs. 2.86%, p = 0.018) decreased with an increase in the CD34+CD133+ progenitor cell quartiles (**Table 1**). The average onset age for AD (p = 0.007) and dementia (p = 0.005) slightly decreased as CD34+CD133+ quartiles increased. Cox proportional hazards regression models were used to study the relationship between CD34+CD133+ cell frequency and AD risk or all-cause dementia risk after adjusting for the covariates (**Table 2**). We found that CD34+CD133+ progenitor cell frequency (log-transformed) was negatively associated with the risk of AD or dementia (Model 1), and this association remained significant after adjusting for covariates, including age, sex, years of education (Model 2), and the addition of *APOE* ε4 (Model 3), showing that a higher frequency of circulating CD34+CD133+ progenitor cells decreased AD risk (HR = 0.67, 95% CI = 0.46-0.96, p = 0.03) and dementia risk (HR = 0.65, 95% CI = 0.48-0.90, p = 0.008). Furthermore, Kaplan‒Meier survival curves based on four CD34+CD133+ cell quartiles revealed that there were significant differences in AD-free probability among the four CD34+CD133+ progenitor cell quartiles (p = 0.034), thus demonstrating a dose-dependent relationship between higher CD34+CD133+ endothelial progenitor cell counts and reduced AD incidence (**Figure 1**).

**Figure 1:**
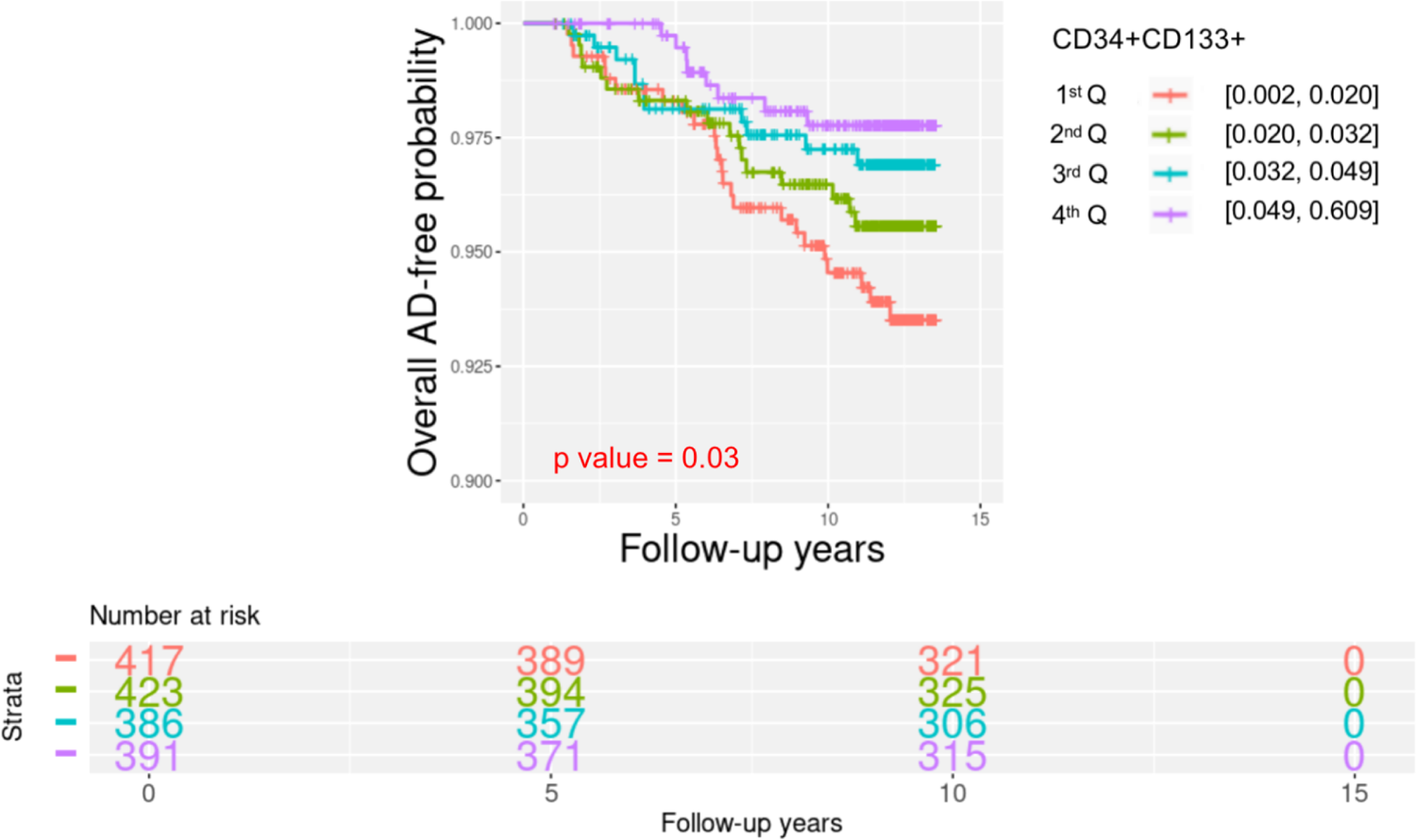
Survival analyses of circulating CD34+CD133+ cells for Alzheimer’s disease risk Circulating CD34+CD133+ endothelial progenitor cells were divided into four quartiles based on the cell numbers (Q1=0.002-0.021; Q2=0.021-0.033; Q3=0.033-0.050; and Q4=0.050-0.609). Kaplan‒Meier survival analysis was used to evaluate the survival-free time before AD over 14 years of follow-up to examine the relationship between CD34+CD133+ quartiles and AD incidence for all of the participants.

**Table 2.**
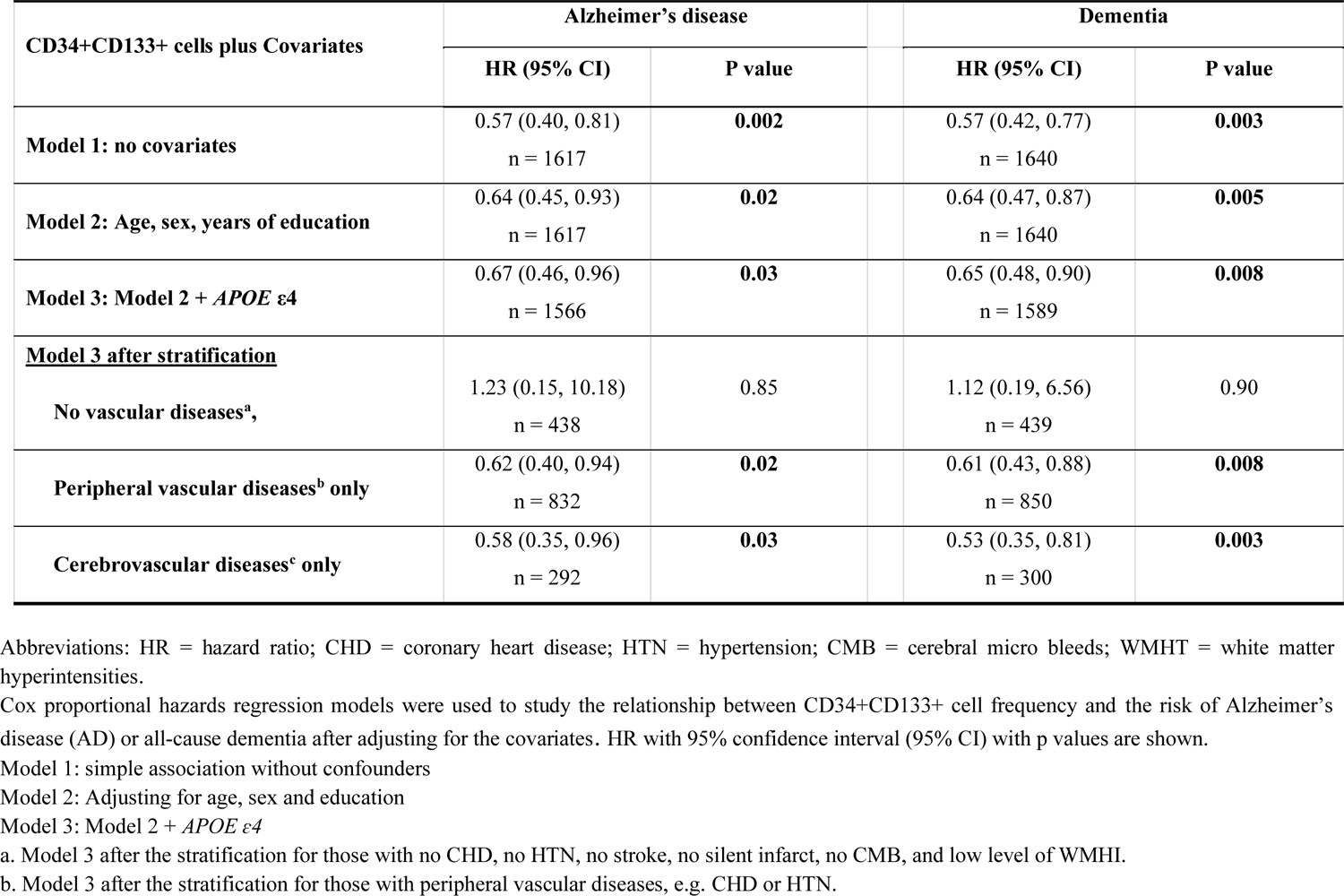
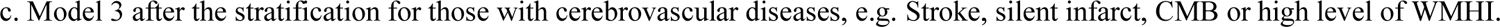
Examination of the relationship between CD34+CD133+ (log transformed) and the risk of AD/dementia risk using Cox proportional hazards regression models after adjusting for different covariates

| CD34+CD133+ cells plus Covariates | Alzheimer's disease |  |  | Dementia |  |
| --- | --- | --- | --- | --- | --- |
|  | HR (95% CI) | P value |  | HR (95% CI) | P value |
| **Model 1: no covariates** | 0.57 (0.40, 0.81)<br>n = 1617 | **0.002** |  | 0.57 (0.42, 0.77)<br>n = 1640 | **0.003** |
| **Model 2: Age, sex, years of education** | 0.64 (0.45, 0.93)<br>n = 1617 | **0.02** |  | 0.64 (0.47, 0.87)<br>n = 1640 | **0.005** |
| **Model 3: Model 2 + *APOE* ε4** | 0.67 (0.46, 0.96)<br>n = 1566 | **0.03** |  | 0.65 (0.48, 0.90)<br>n = 1589 | **0.008** |
| **<u>Model 3 after stratification</u>** |  |  |  |  |  |
| **No vascular diseases<sup>a</sup>,** | 1.23 (0.15, 10.18)<br>n = 438 | 0.85 |  | 1.12 (0.19, 6.56)<br>n = 439 | 0.90 |
| **Peripheral vascular diseases<sup>b</sup> only** | 0.62 (0.40, 0.94)<br>n = 832 | **0.02** |  | 0.61 (0.43, 0.88)<br>n = 850 | **0.008** |
| **Cerebrovascular diseases<sup>c</sup> only** | 0.58 (0.35, 0.96)<br>n = 292 | **0.03** |  | 0.53 (0.35, 0.81)<br>n = 300 | **0.003** |

Abbreviations: HR = hazard ratio; CHD = coronary heart disease; HTN = hypertension; CMB = cerebral micro bleeds; WMHT = white matter hyperintensities.
Cox proportional hazards regression models were used to study the relationship between CD34+CD133+ cell frequency and the risk of Alzheimer's disease (AD) or all-cause dementia after adjusting for the covariates. HR with 95% confidence interval (95% CI) with p values are shown.
Model 1: simple association without confounders
Model 2: Adjusting for age, sex and education
Model 3: Model 2 + *APOE* ε4
a. Model 3 after the stratification for those with no CHD, no HTN, no stroke, no silent infarct, no CMB, and low level of WMHI.
b. Model 3 after the stratification for those with peripheral vascular diseases, e.g. CHD or HTN.
c. Model 3 after the stratification for those with cerebrovascular diseases, e.g. Stroke, silent infarct, CMB or high level of WMHl.

Cox proportional hazards regression models were also used to examine the relationships between other progenitor cells, including CD34+, CD34+CD133-, CD34-CD133+, CD34-CD133-, or CD34+KDR+(VEGFR2) cells, and AD dementia risk. In addition, we studied mature cells including CD31+, CD31-, CD31+DIM, and CD31+lymphoid cells for AD risk using the same analyses. Unlike CD34+CD133+ progenitor cells, none of these cell types impacted the risk of AD dementia (**Supplement Table 1**).

### The relationship between CD34+CD133+ endothelial progenitor cells and reduced AD/dementia risk in the presence of peripheral and central vascular diseases

As expected, compared to those participants without any vascular diseases (0.78%), participants with peripheral vascular diseases, including coronary heart disease (CHD) (8.27%, p = 4.1e-05) and HTN (7.42%, p = 7.5e-05), and participants with cerebrovascular diseases, including clinical diagnoses of stroke (10.83%, p = 3.3e-06), silent infarct (10.97%, p = 1.8e-06), CMB (15.12%, p = 1.8e-08), and high white matter hyperintensities (WMHI) (7.59%, p = 7.0e-05), had significantly higher rates of incident AD (**Figure 2A**). A similar but stronger trend was observed for the incident rates of all-cause dementia in participants with these vascular diseases, although this can be primarily attributed to the fact that there were more cases of dementia than AD (**Figure 2B**). When each peripheral vascular or cerebrovascular disease was added as a covariate into the cox proportion regression model, in addition to adjusting for age, sex, years of education and *APOE* ε4 (Model 3 plus), the negative relationship between CD34+CD133+ progenitor cells and the risk of AD dementia remained (**Supplement Table 2**).

**Figure 2.**
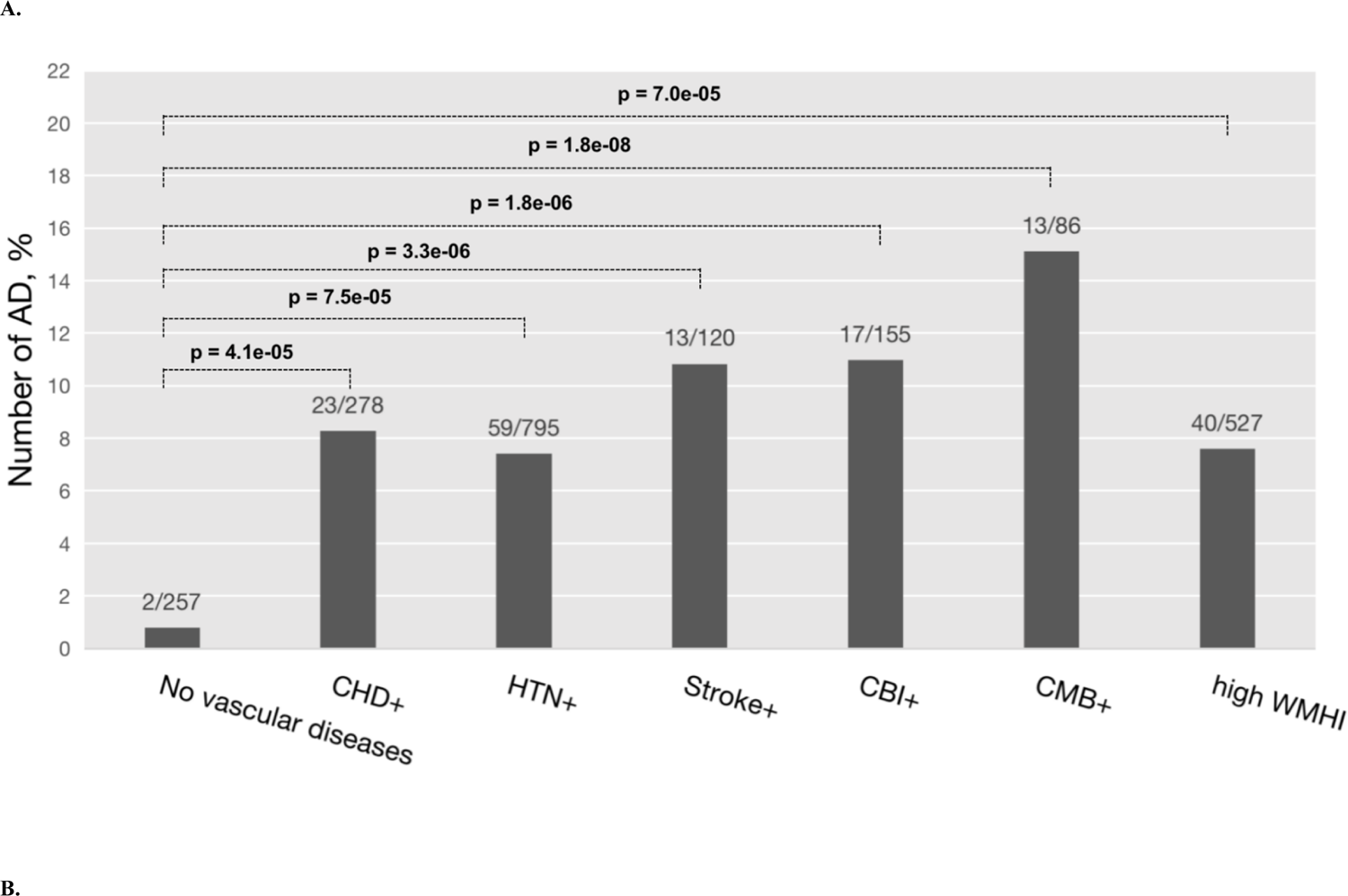
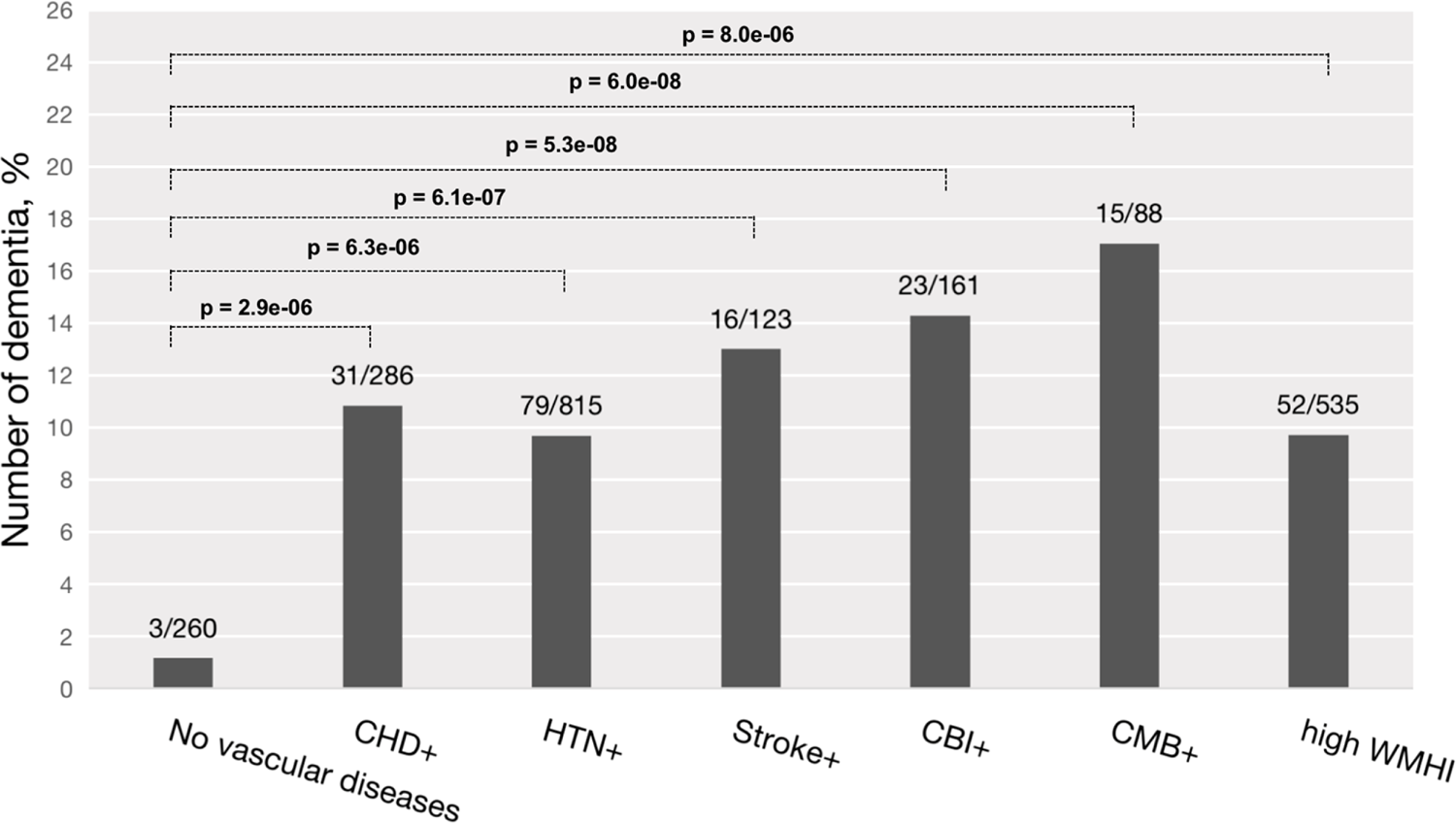
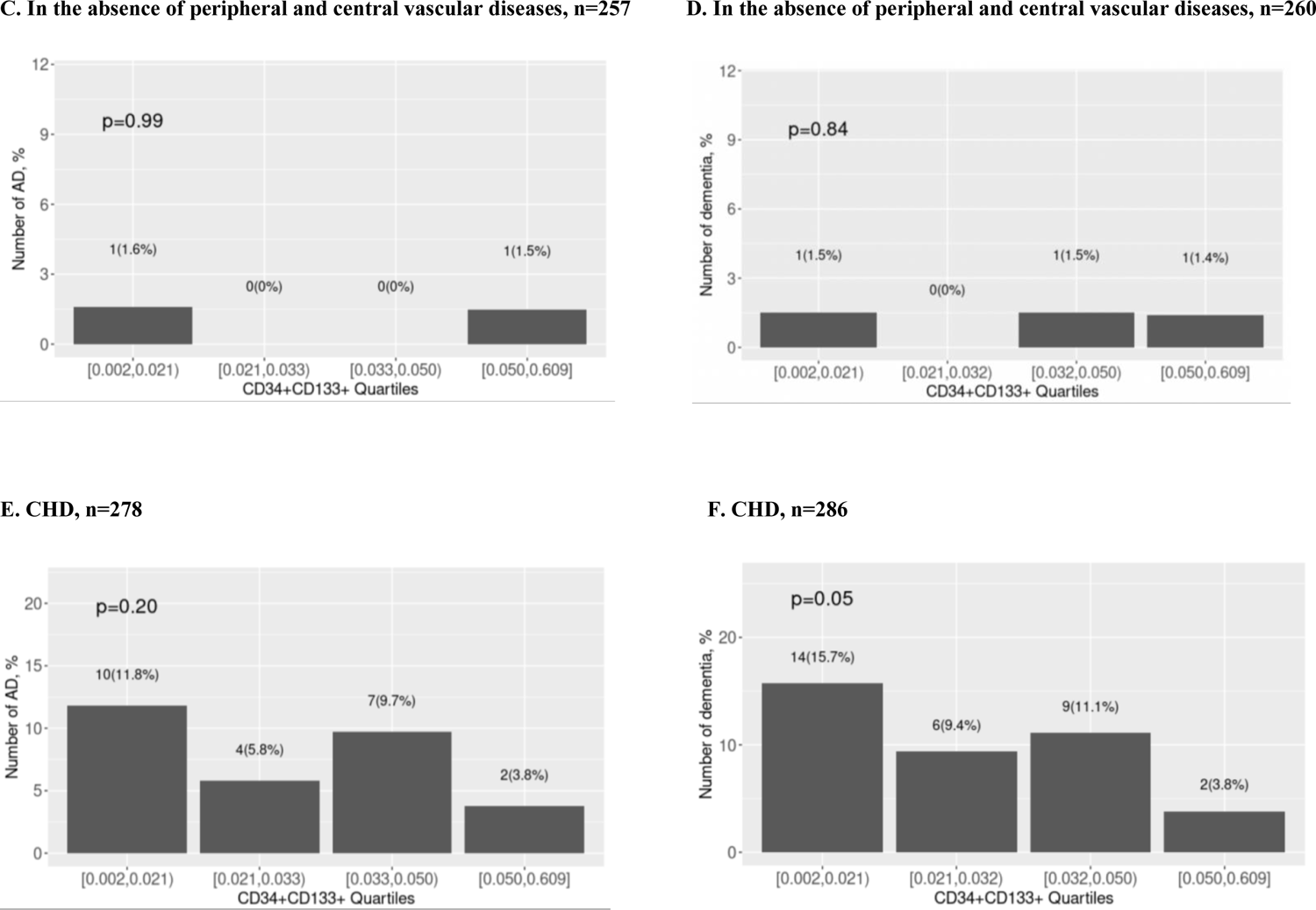
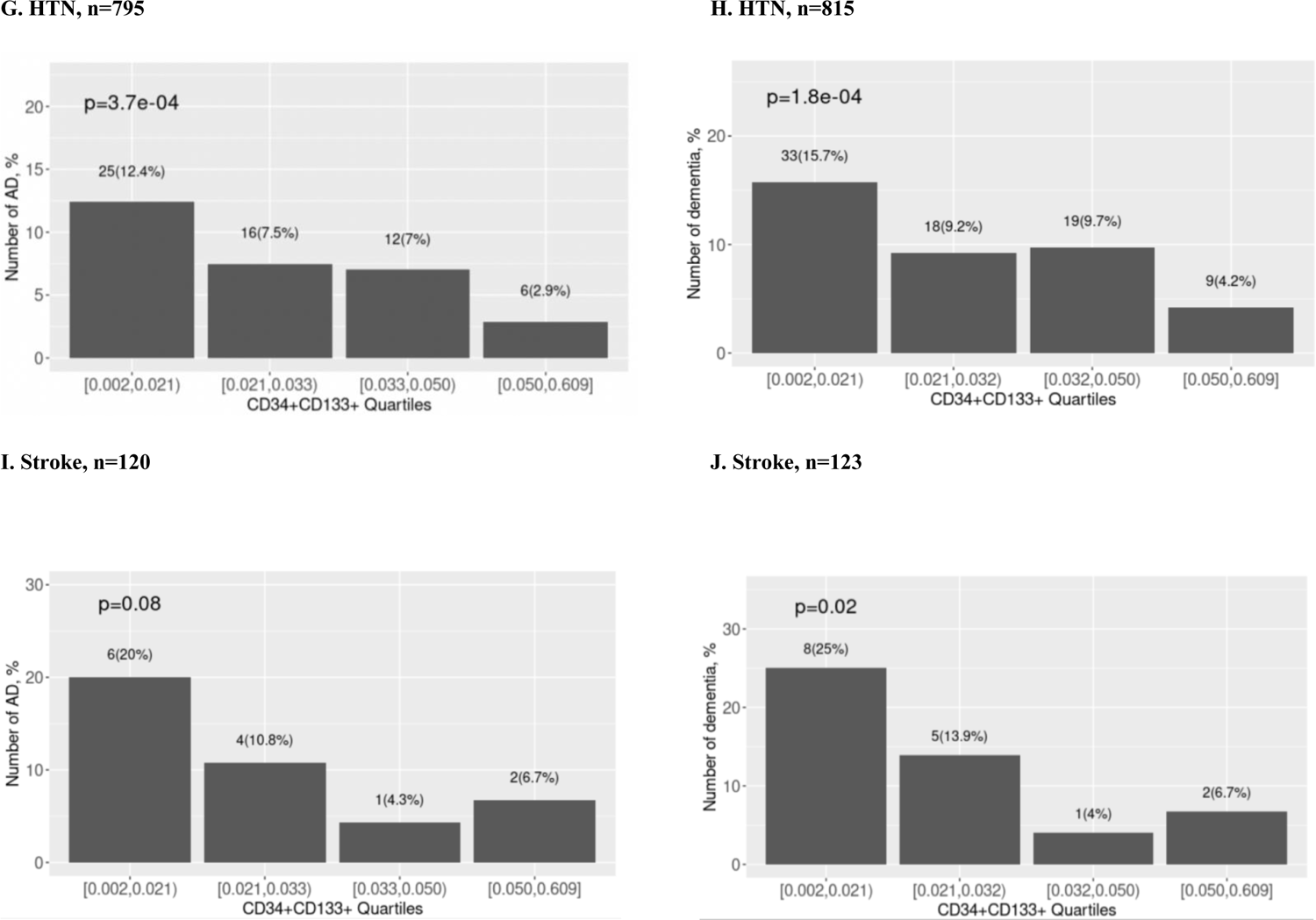
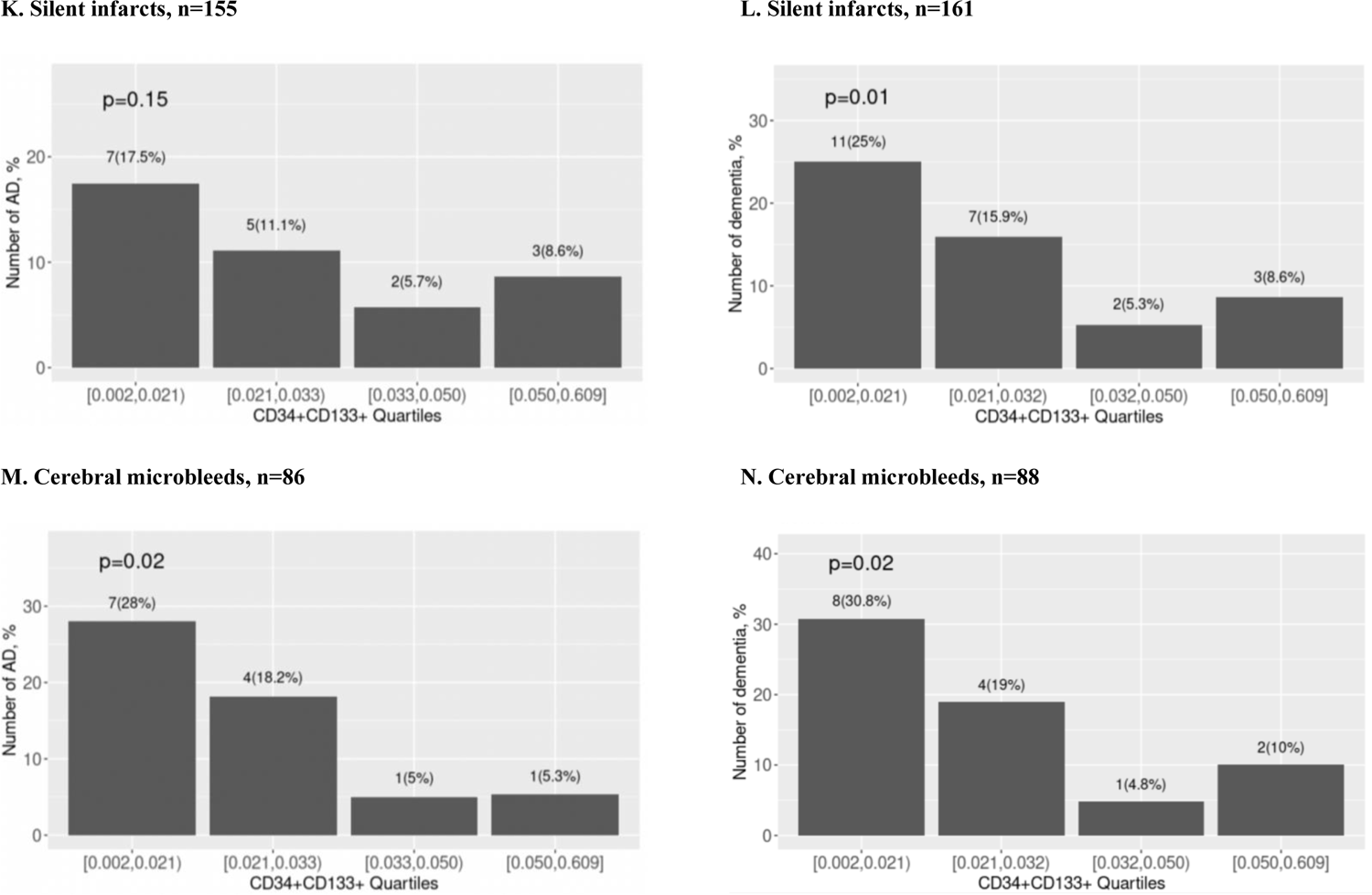
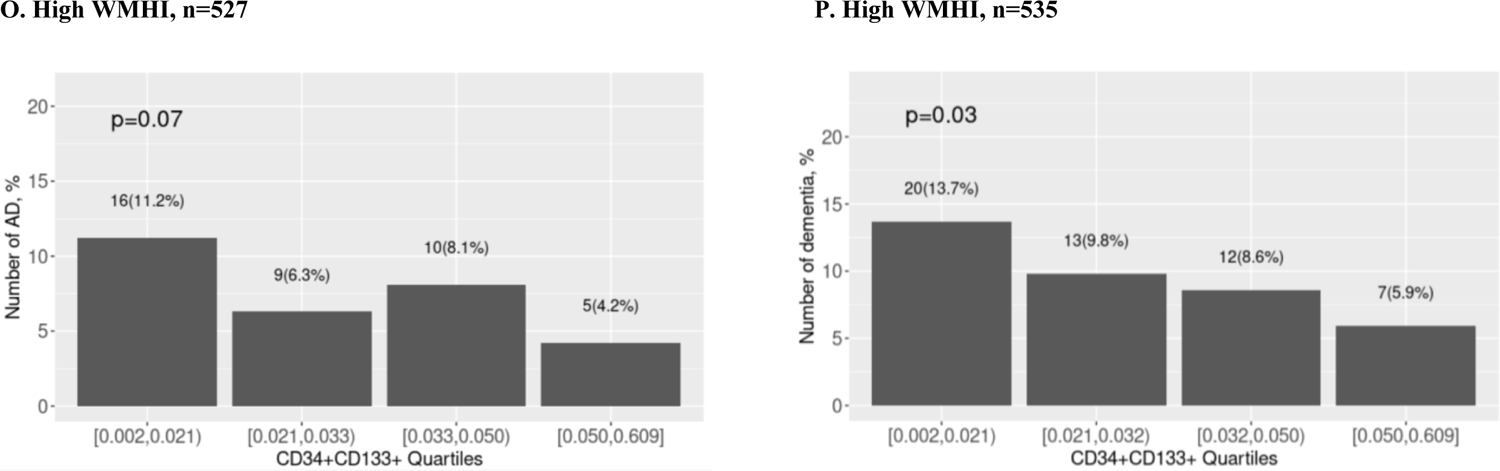
Circulating CD34+CD133+ progenitor cells decrease AD dementia risk in the presence of peripheral and central vascular diseases Participants were stratified into those individuals with the absence or presence of different vascular diseases, and the chi-squared test was used with the indicated *p* values to examine the numbers and percentages of incident AD (**A**) and dementia (**B**). Participants were also stratified into the absence of any peripheral and central vascular diseases (**C, D**), CHD = Coronary heart disease (**E, F**), HTN = hypertension (**G, H**), stroke (**I, J**), silent infarcts (**K, L**), cerebral microbleeds (**M, N**), and high WMHI = white matter hyperintensities (**O, P**); they were further divided based on CD34+CD133+ endothelial progenitor cell quartiles, followed by the use of the chi-squared test with *p* values to examine incident AD (**C, E, G, I, K, and M**) and dementia (**D, F, H, J, L, N, and P**).

We further hypothesized that endothelial progenitor cells would play roles in reducing AD risk especially in the presence of vascular pathologies when endothelial cells are damaged. Indeed, after stratification, whereas this trend was not observed in individuals without any vascular diseases, increased CD34+CD133+ endothelial progenitor cell frequency was significantly associated with decreased AD risk among participants who had peripheral vascular diseases (HR = 0.62, 95% CI: 0.40-0.94, p = 0.02) or cerebrovascular disease (HR = 0.58, 95% CI: 0.35-0.96, p = 0.03) (**Table 2**). Similar trends were observed for all-cause dementia risk among those individuals with peripheral vascular diseases (HR = 0.61, 95% CI: 0.43-0.88, p = 0.008) or with cerebrovascular disease (HR = 0.53, 95% CI: 0.35-0.84, p = 0.003). Consistently, in the absence of any vascular disease, increased CD34+CD133+ frequency quartiles were not associated with a change in AD or dementia incidence (**Figure 2C and 2D**). In contrast, CD34+CD133+ progenitor cell quartiles significantly reduced AD risk in the presence of vascular diseases in a dose-dependent pattern (**Figure 2E-P**). Specifically, increased CD34+CD133+ progenitor cell quartiles had a significantly dose-dependent association with decreased AD risk among participants who had HTN (p=3.7e-04) (**Figure 2G**) and CMB (p=0.02) (**Figure 2M**). Regarding dementia risk, similar trends for increasing CD34+CD133+progenitor cell quartiles to reduce dementia risk were observed in different vascular diseases in addition to HTN and CMB, specifically among those with CHD (p=0.05) (**Figure 2F**), HTN (p=1.8e-04) (**Figure 2H**), stroke (p=0.02) (**Figure 2J**), silent infarct (p=0.01) (**Figure 2L**), CMB (p=0.02) (**Figure 2N**) or in a high WMHI level (p=0.03) (**Figure 2P**).

### Interaction effect between KIRREL3 *or* EXOC6B *homozygous genotype* and CD34+CD133+ endothelial progenitor cells on AD risk

Studies have shown that brain endothelial damage increases AD pathogenesis in the brain among *APOE* ε4 carriers,^4^ therefore, we hypothesized that peripheral circulating CD34+CD133+ endothelial progenitor cells can further reduce AD risk in persons with particular genotypes. To test this hypothesis, we first conducted a genome-wide association study (GWAS) for AD with the interaction term between CD34+CD133+ cell frequency and single nucleotide polymorphism (SNP) as predictor using a logistic regression model. For SNPs passing a genome-wide significance (p < 5.0e-08), a Cox regression model was applied to validate the interaction effects on AD incidence. As depicted in the Manhattan plot, 8 SNPs showed significant interactions with CD34+CD133+ frequency for AD risk (**Figure 3A**). Seven of the 8 SNPs with a p value <5.0×10^-8^ were located in gene *KIRREL3* on chromosome 11 (two of the 7 SNPs selected for further analysis: rs580382 and rs4144611) and 1 of the 8 SNPs was in gene *EXOC6B* (chromosome 2, rs61619102) (**Figure 3A, Supplement Figure 2**). These associations are supported by evidence with surrounding SNPs (**Figures 3B and 3C, Supplement Table 3**). Notably, for these 3 SNPs, although their main SNP effects were not associated with AD risk (p>0.40), their interactive effects with CD34+CD133 cells were significantly associated with AD after adjusting for age, sex, years of education, *APOE* ε4, and principle components (PCs) using both logistic and Cox regression analyses (**Table 3**).

**Figure 3.**
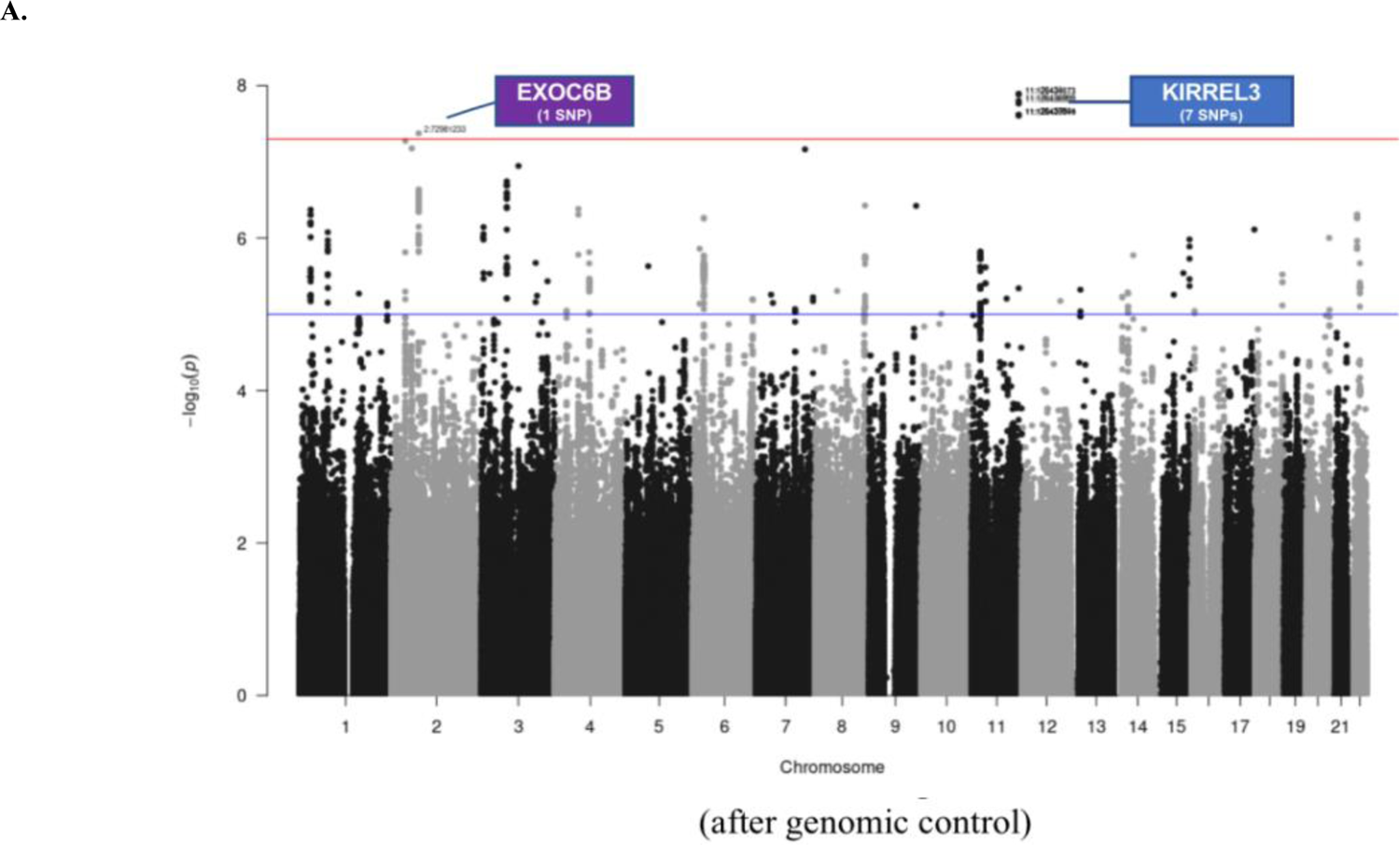
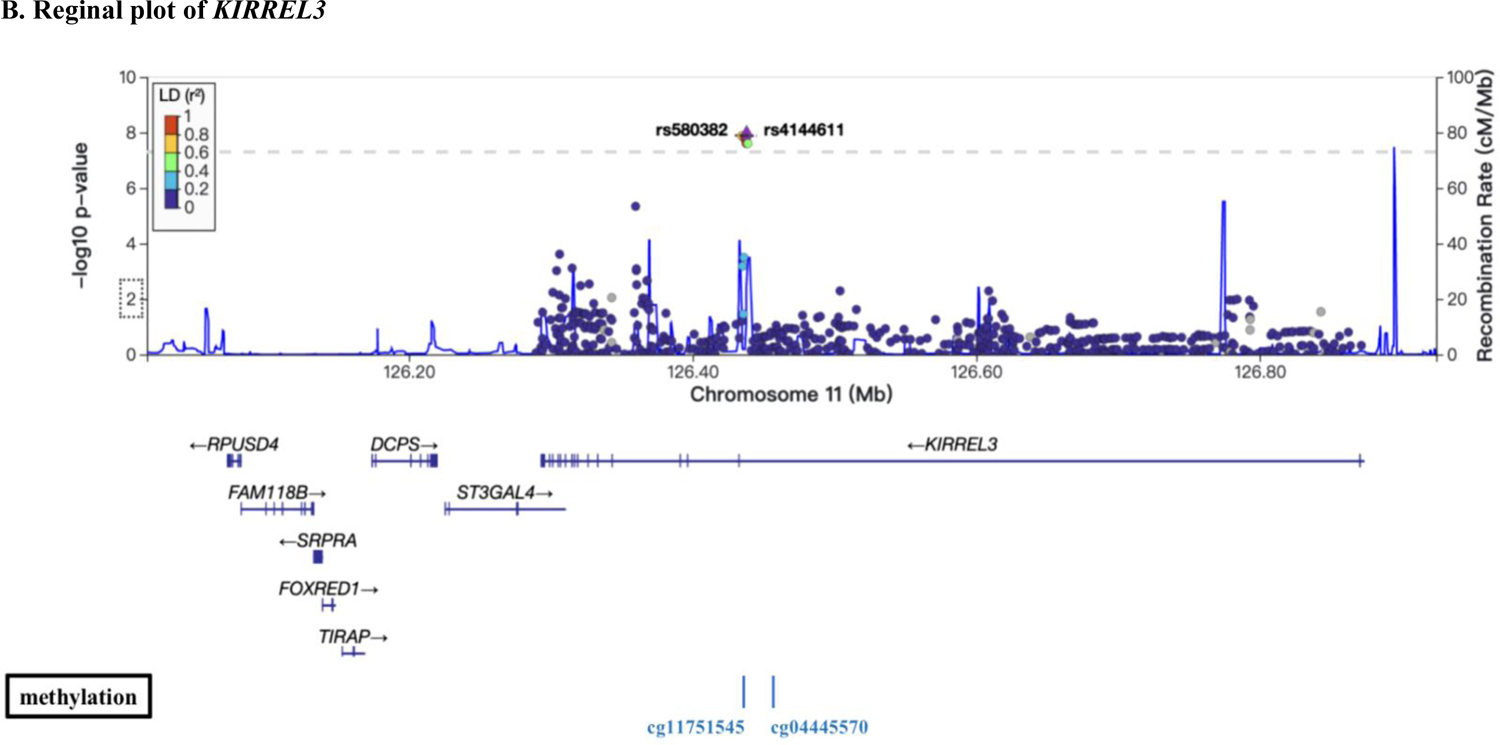
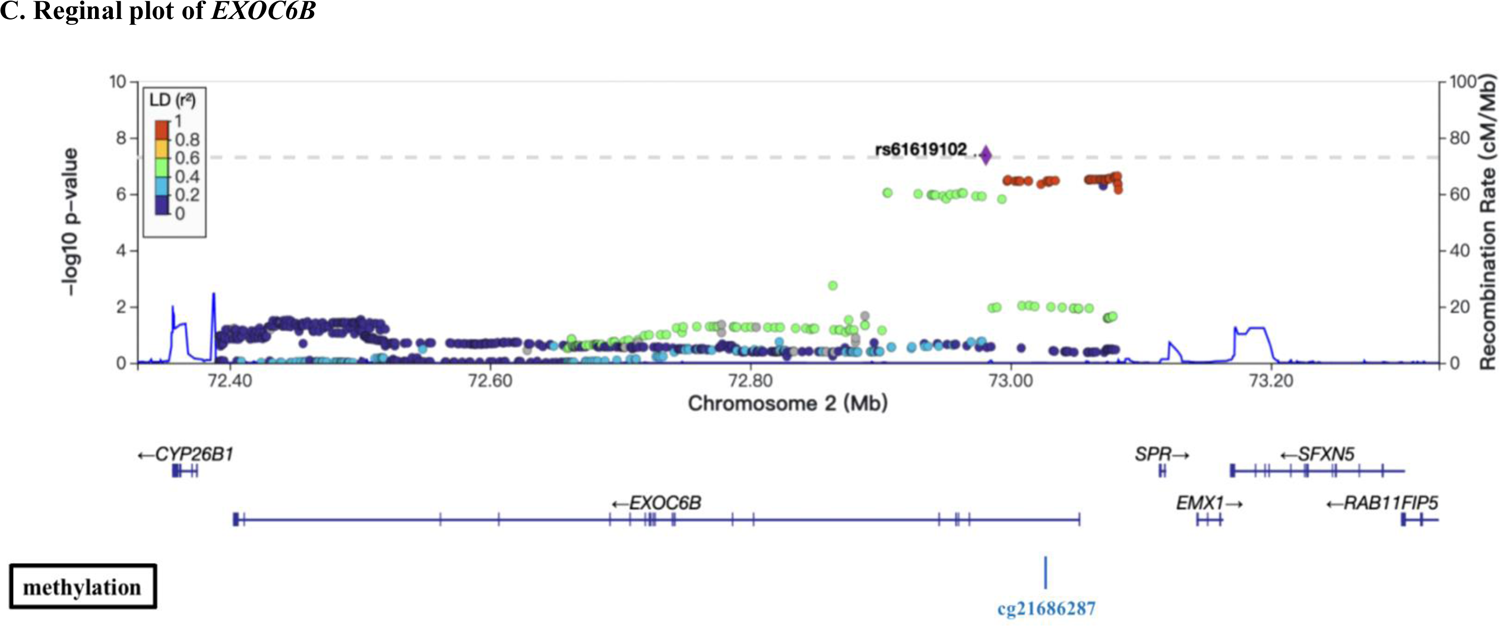
GWAS on the interactive effects of genetic factors and circulating CD34+CD133+ cells on Alzheimer’s disease risk The FSH analytical pipeline was used to perform genome-wide association studies (GWAS). The interaction between genome-wide SNP dosage and circulating CD34+CD133+ cells for AD risk was tested by using the GEEPACK model (logistic regression utilizing generalized estimating equations) adjusting for age, sex, years of education, and the first 10 PCs. As shown in the Manhattan plot, 8 SNPs showed significant interactions with CD34+CD133+ frequency on AD risk (**A**). Seven of the identified SNPs with a p value ≤ 5*10^-8^ originated from gene Kirre Like Nephrin Family Adhesion Molecule 3 (*KIRREL3*) on chromosome 11 (rs580382 and rs4144611 were selected for further analysis), and 1 of them originated from Exocyst Complex Component 6B (*EXOC6B*) (chromosome 2, rs61619102). Only r^2^ ≥ 0.1 and SNPs with minor allele frequency (MAF) ≥10% were considered. Manhattan plot (**A**) used for visualization to identify the genotypes of two genes (*KIRREL3* and *EXOC6B*) that had interactive effects with CD34+CD133+ endothelial progenitor cells for AD risk. Gene structures and surrounding regions of *KIRREL3* (**C**) and *EXOC6B* (**D**) are presented by using LocusZoom.

**Table 3.**
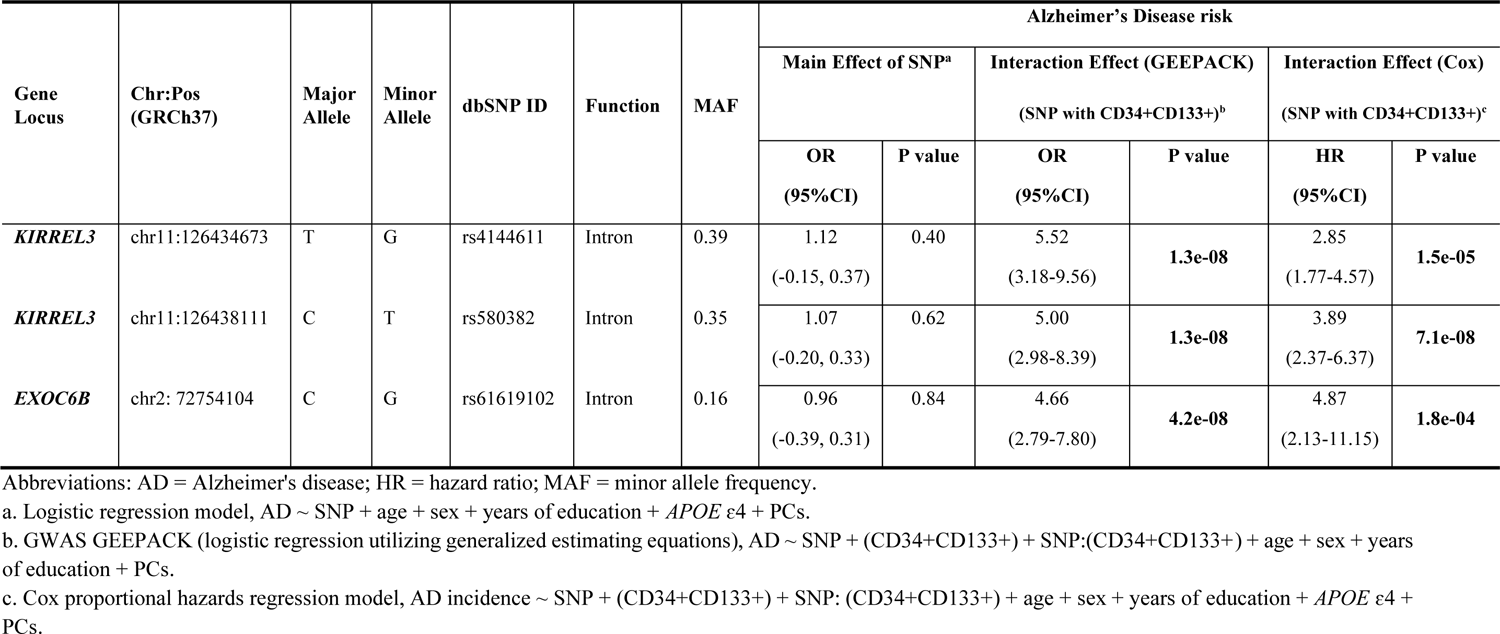
Interactions between SNPs of *KIRREL3* or *EXOC6B* and CD34+CD133+ on AD risk in FHS

Consistently, in genotype stratification analysis adjusting for age, sex, years of education, *APOE* ε4, and PCs, we found that CD34+CD133+ frequency (log-transformed and continuous) was negatively associated with AD incidences among the homozygotes of *KIRREL3* rs4144611 TT carriers (HR = 0.29, 95% CI: 0.15-0.57, p<0.001), *KIRREL3* rs580382 CC carriers (HR = 0.31, 95% CI: 0.17-0.57, p<0.001) and *EXOC6B* rs61619102 CC carriers (HR = 0.49, 95% CI: 0.31-0.75, p<0.001) (**Supplementary Table 4**). Further examination of these findings revealed a dose-dependent effect of increased CD34+CD133+ cell quartiles on the reduction in AD risk among *KIRREL3* rs4144611 TT or rs580382 CC homozygotes as well as in *EXOC6B* rs61619102 CC homozygotes (**Table 4, Supplementary Table 5**). In contrast, these impacts from CD34+CD133+ cells were not present in the counterpart genotypes of these 3 SNPs. We further tested each genotype e.g. *KIRREL3* rs4144611 either TG or GG; rs580382 either CT or TT; as well as *EXOC6B* rs61619102 either GC or GG carriers, using Cox regression model and found that none of them had interactive effects with CD34+CD133+ progenitor cells on AD risk (**Supplementary Table 5**).

**Table 4.** Circulating CD34+CD133+ cells for Alzheimer’s disease in the context of genetic background in FHS, adjusting for age, sex, years of education, *APOE* ɛ4, and PCs^a^

| rs4144611 (*KIRREL3*) |  |  |  | rs580382 (*KIRREL3*) |  |  |  | rs61619102 (*EXOC6B*) |  |  |  |
| --- | --- | --- | --- | --- | --- | --- | --- | --- | --- | --- | --- |
| genotype | CD34+CD133+<br>cutoffs | HR<br>(95% CI) | P value | genotype | CD34+CD133+<br>cutoffs | HR<br>(95% CI) | P value | genotype | CD34+CD133+<br>cutoffs | HR<br>(95% CI) | P value |
| **TT** | 25% | 0.18<br>(0.07-0.46) | **3.3e-04** | **CC** | 25% | 0.21<br>(0.09-0.47) | **1.7e-04** | **CC** | 25% | 0.48<br>(0.26-0.88) | **0.02** |
|  | 50% | 0.11<br>(0.03-0.42) | **0.001** |  | 50% | 0.16<br>(0.05-0.47) | **9.6e-04** |  | 50% | 0.38<br>(0.19-0.74) | **0.005** |
|  | 75% | *N/A*<br>(Zero AD) | **0.006<sup>a</sup>** |  | 75% | *N/A*<br>(Zero AD) | **0.002<sup>a</sup>** |  | 75% | 0.25<br>(0.08-0.81) | **0.02** |
| **GG+TG** | 25% | 1.02<br>(0.46-2.25) | 0.96 | **TT+CT** | 25% | 1.80<br>(0.68-4.82) | 0.24 | **GG+GC** | 25% | 2.22<br>(0.39-12.64) | 0.37 |
|  | 50% | 1.54<br>(0.76-3.15) | 0.23 |  | 50% | 2.44<br>(1.07-5.56) | **0.03** |  | 50% | 12.38<br>(2.00-76.65) | **0.007** |
|  | 75% | 1.39<br>(0.64-3.05) | 0.41 |  | 75% | 1.80<br>(0.80-4.07) | 0.16 |  | 75% | 5.68<br>(1.65-19.61) | **0.006** |

Participants were first stratified into *KIRREL3* rs4144611 TT vs. GG+TG; rs580382 CC vs. TT+CT as well as in *EXOC6B* rs61619102 CC vs. GG+GC genotype groups. Cox proportional hazards regression model, AD incidence ~ (CD34+CD133+ cutoffs: 25%, 50% and 75%) + age + sex + years of education + *APOE* ε4 + PCs was used for each genotype group.
a. When CD34+CD133+ is higher than the 75% percentile, there are no AD cases among these genotypes, *KIRREL3* rs4144611 TT, *KIRREL3* rs580382 CC and *EXOC6B* rs61619102 CC; thus, log-rank statistics are instead used to compare groups.

These relationships were also observed in Kaplan‒Meier survival curves. Among the homozygotes of *KIRREL3* rs4144611 TT carriers (p=0.003, **Figure 4A**), *KIRREL3* rs580382 CC carriers (p=1.2e-04, **Figure 4B**), and *EXOC6B* rs61619102 CC carriers (p=0.001, **Figure 4C**), higher CD34+CD133+ frequency quartiles corresponded to a lower incidence of AD development and difference in AD-free probability with statistical significance. Again, among their counterpart genotype carriers (*KIRREL3* rs4144611 GG+GT or rs580382 TT+CT carriers and *EXOC6B* rs61619102 GG+GC carriers), there were no such relationships between CD34+CD133+ quartiles and AD or all-cause dementia risk. Additionally, unlike for AD dementia risk, we did not find associations of these genotypes or the interactive effects of these genotypes and CD34+CD133+ cells for any vascular disease in **Table 1** (data not shown).

**Figure 4:**
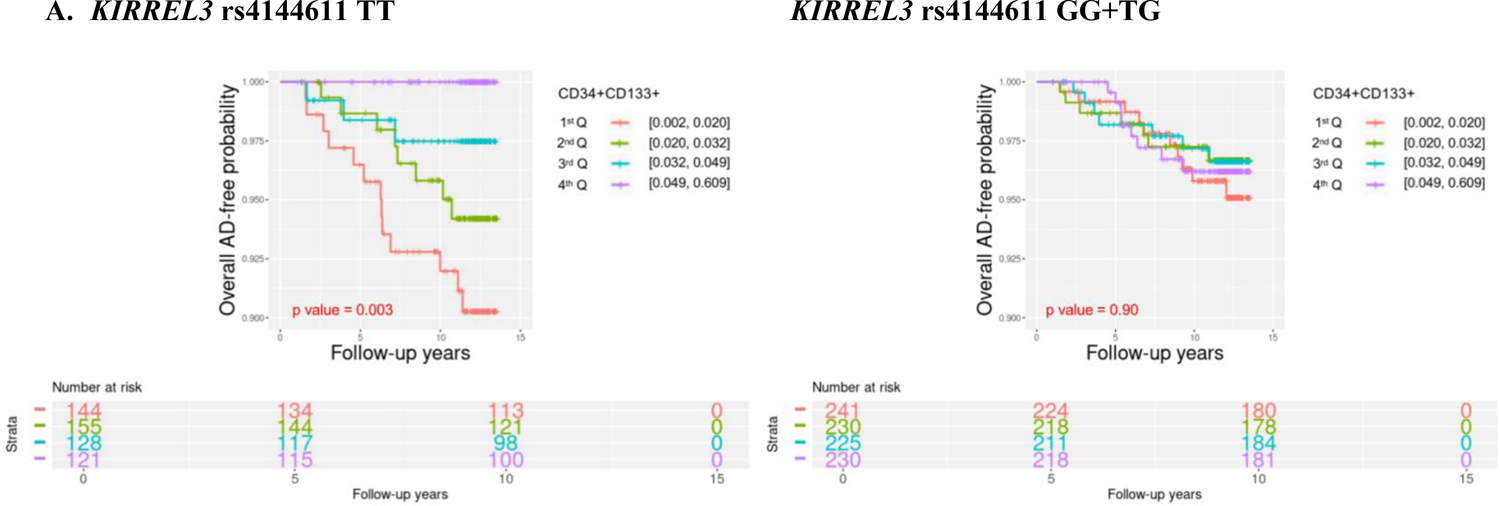
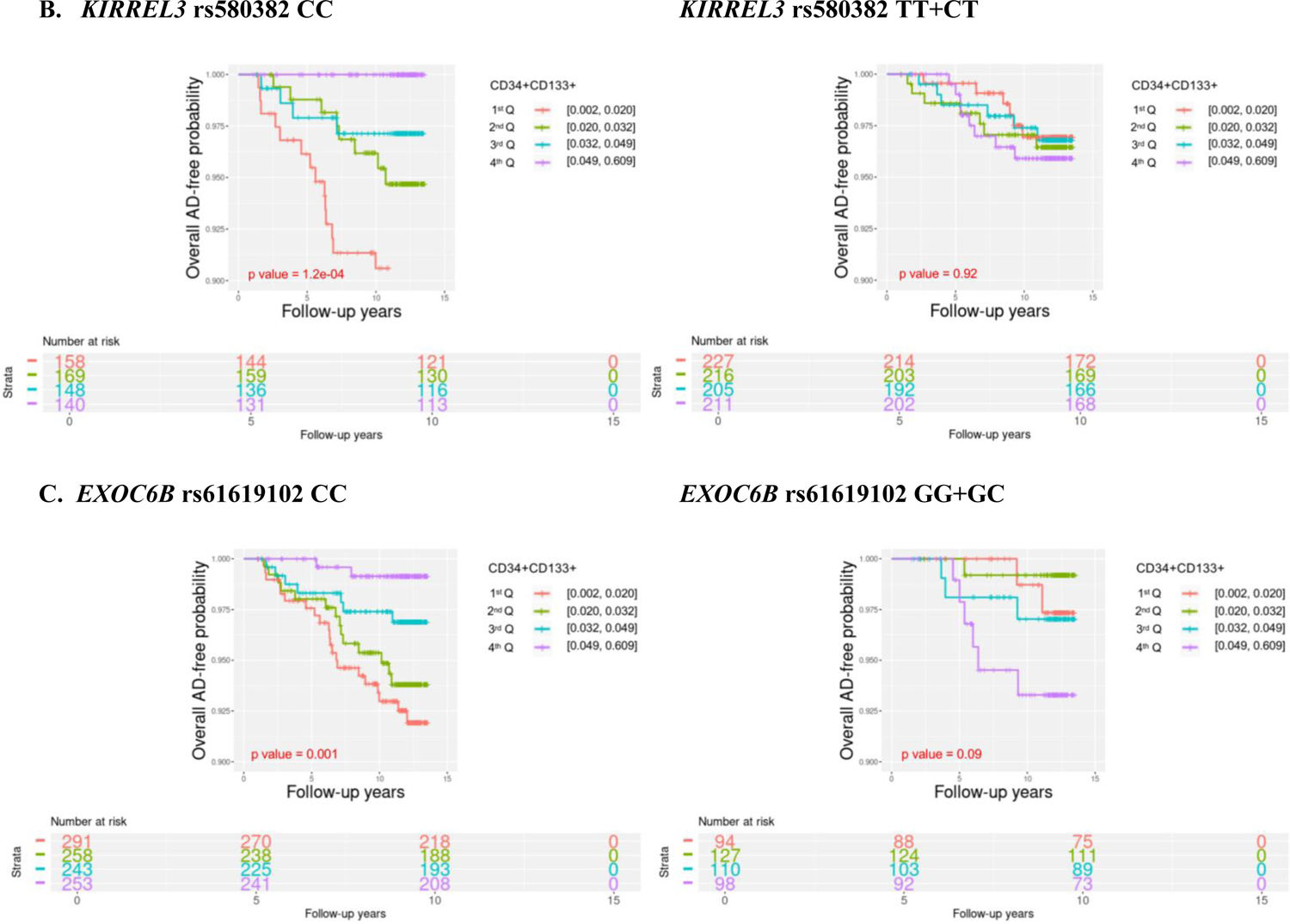
Survival analyses of circulating CD34+CD133+ cells for Alzheimer’s disease risk in the context of genetic background Circulating CD34+CD133+ endothelial progenitor cells were divided into four quartile groups based on the cell numbers (Q1=0.002-0.021; Q2=0.021-0.033; Q3=0.033-0.050; and Q4=0.050-0.609). Kaplan‒Meier survival analysis was used to evaluate the survival-free time before AD over 14 years of follow-up to examine the relationship between CD34+CD133+ quartiles and AD incidence for participants stratified based on the following genotypes: *KIRREL3* rs4144611 major homozygotes TT vs. GG+TG (**A**), *KIRREL3* rs580382 major homozygotes CC vs. TT+CT (**B**), and *EXOC6B* rs61619102 major homozygotes CC vs. GG+GC (**C**).

### Exploration of KIRREL3 and EXOC6B genes in Alzheimer’s disease

To address why these two genes could interact with CD34+CD133+ endothelial progenitor cells for AD risk, we explored if the mRNA expressions level of gene *KIRREL3* and *EXOC6B* are associated with AD pathology. We first used the RNAseq dataset generated from the dorsolateral prefrontal cortex (DLPFC) region and monocytes in the ROSMAP study. As shown in **Figure 5A-a**, brain *KIRREL3* expression was associated with AD (p=0.02), but peripheral *KIRREL3* expression was not. Compared to controls, brain expression level of *KIRREL3* was lower in AD brains specifically in the temporal cortex (TCX) (p = 2.7e-05) regions (**Figure 5A-b**). To investigate the relationship between *KIRREL3* SNPs and lower mRNA levels in the AD brain, we investigated the epigenetics of *KIRREL3* SNPs and found that the methylation site cg11751545 was consistently negatively correlated with *KIRREL3* gene expression in DLPFC (**Supplement Figure 2A**). Two *KIRREL3* genotypes (rs4144611 GG+GT or rs580382 TT+CT) were negatively associated with DNA methylation level of two CpG sites within *KIRREL3* (cg11751545; cg04445570; p<0.05) (**Figure 3B, Supplement Figure 2B**). Based on these results, we hypothesized that the rescue effect of CD34+CD133+ progenitor cells may reduce AD risk only in vulnerable *KIRREL3* rs4144611 TT carriers (**Table 4** and **Figure 4A**) and rs580382 CC carriers (**Table 4; Figure 4B** and **Figure 5A-c**) because the low brain expression of *KIRREL3* is regulated by the high brain methylation of cg11751545 in these carriers.

**Figure 5.**
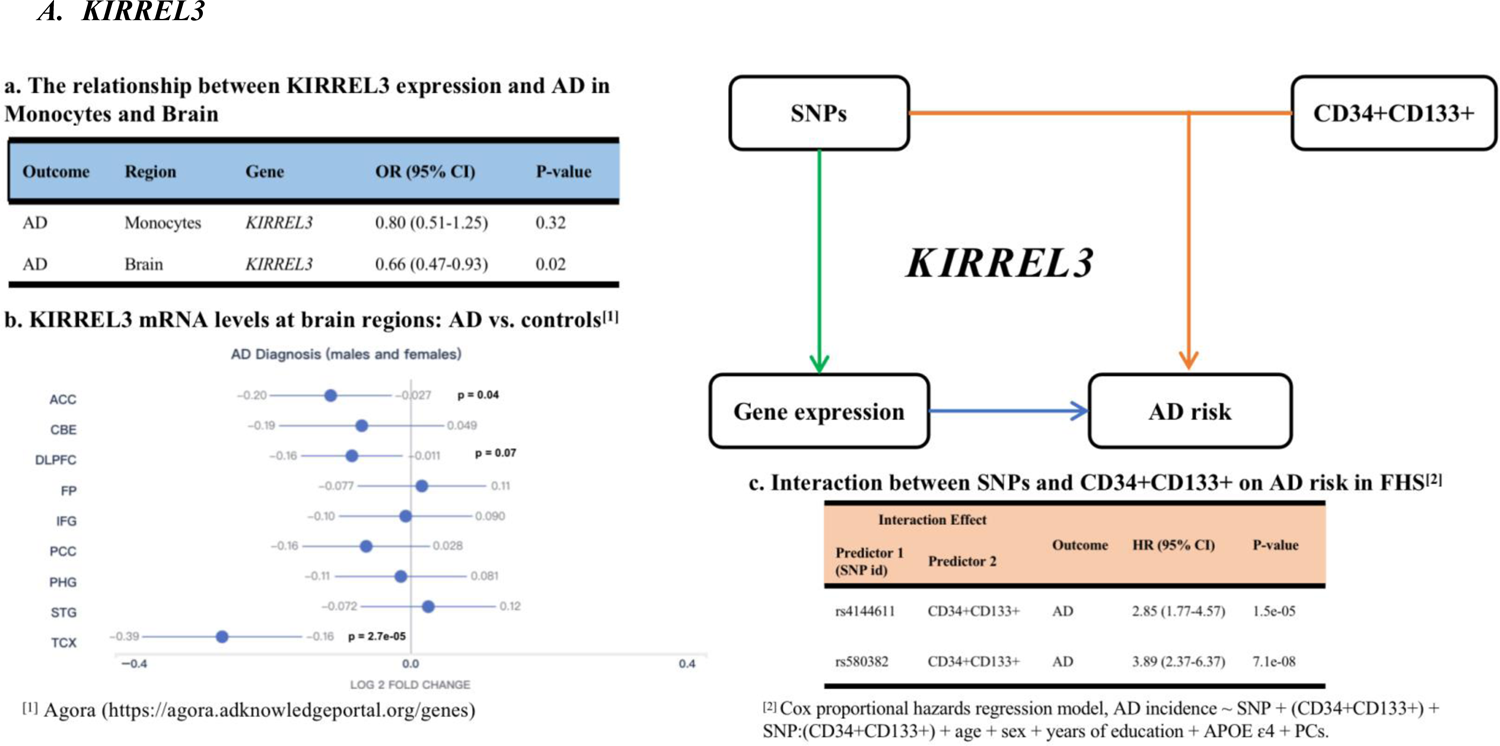
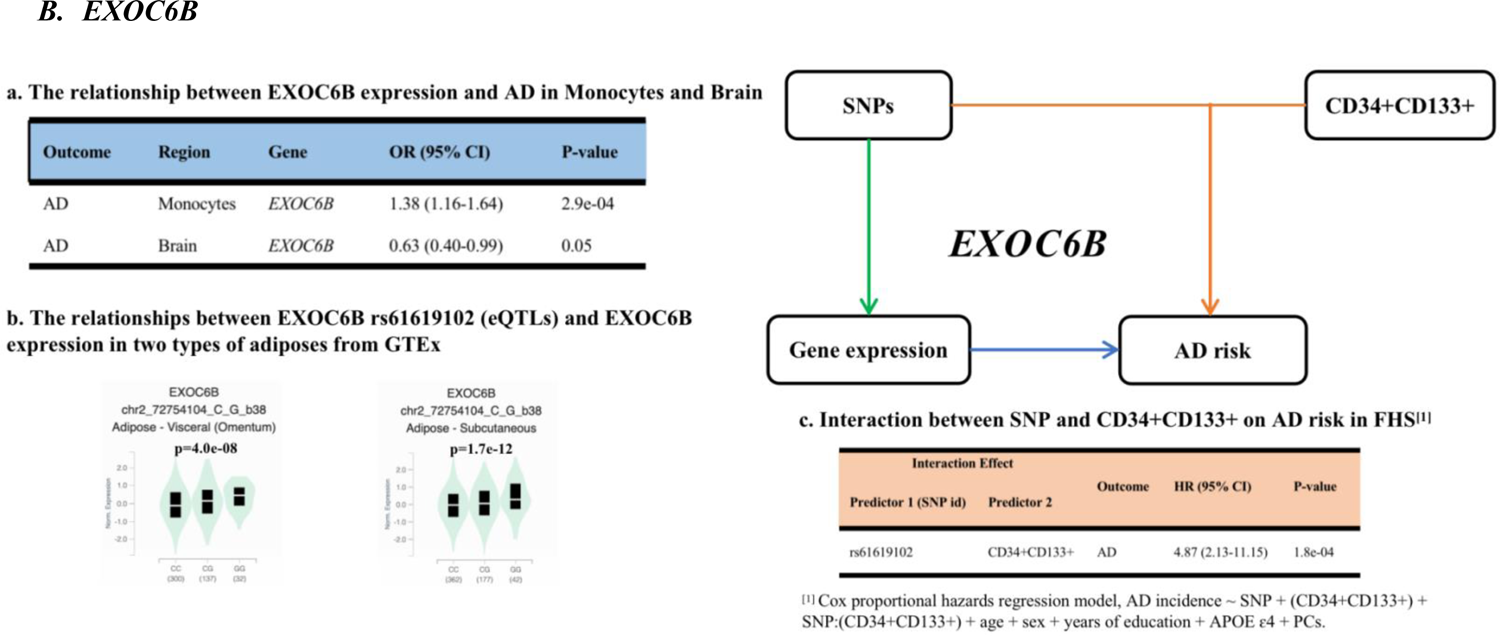
The relationships between the SNPs, gene expression, and Alzheimer’s disease We hypothesized that the gene expression levels of *KIRREL3* (**A**) and *EXOC6B* (**B**) are involved in AD pathology, thus leading to their genotypes having interactive effects with circulating CD34+CD133+ endothelial progenitors for AD risk. (A) *KIRREL3*: a. by using the ROSMAP dataset and logistic regression, the relationships between *KIRREL3* expression in monocytes and brain and AD pathology are shown; b. *KIRREL3* mRNA levels across brain regions were compared between AD and controls obtained from Agora; c. interaction between two *KIRREL3* SNPs and CD34+CD133+ on AD risk in FHS. ACC = The anterior cingulate cortex; CBE = cerebellum; DLPF = The dorsolateral prefrontal cortex; FP = The frontal pole; IFG = The inferior frontal gyrus; PCC = The posterior cingulate cortex; PHG = The parahippocampal gyrus; STG = The superior temporal gyrus; TCX = The temporal cortex (B) *EXOC6B*: **a**. by using the ROSMAP dataset and logistic regression, the relationships between *EXOC6B* expression in monocytes and brain and AD pathology are shown, thus indicating that high peripheral expression of *EXOC6B* expression was significantly associated with AD risk; **b**. peripheral eQTL results for the *EXOC6B* genotype in two types of adipose tissues from GTEx are illustrated, thus suggesting that the *EXOC6B* cc allele with low peripheral expression and AD risk can be rescued by circulating CD34+CD133+ endothelial progenitor cells for AD risk; **c**. the interaction between the *EXOC6B* SNP and CD34+CD133+ cells impacted AD risk in FHS.

While peripheral monocyte expression of *EXOC6B* was significantly and positively associated with AD (p < 0.001), the brain expression of *EXOC6B* was not associated with AD after Bonferroni correction (**Figure 5B-a**). Consistently, *EXOC6B* rs61619102 G allele was positively associated with *EXOC6B* gene expression as an eQTL compared to C allele in the following two types of peripheral tissues: visceral and subcutaneous adipose tissues (**Figure 5B-b**). Thus, we reasoned that the impact of *EXOC6B* polymorphisms on AD was mainly elicited through the peripheral system and that the *EXOC6B* rs61619102 homozygous genotype (CC) itself (which was associated with a low level of expression in the peripheral system) interacted with circulating CD34+CD133+ progenitor cells to reduce AD risk (**Table 4; Figure 4C; Figure 5B-c**).

## Discussion

As peripheral and central vascular diseases increase AD risk, it is possible that there are some protective factors *in vivo* antagonizing or rescuing cerebrovascular pathologies for reducing the risk of AD. However, this kind of *in vivo* protection is largely unclear. To the best of our knowledge, this study was the first to illustrate that different vascular pathologies increased the risk of consequential AD development which was depended on two factors: 1) the presence of vasculature damage and 2) the ability to repair/reconstitute dysfunctional endothelia by circulating CD34+CD133+ progenitor cells. As endothelial repair for peripheral blood vessels is mainly performed by circulating endothelial progenitor cells with the co-expression of CD34 and CD133,^12^ circulating CD34+CD133+ progenitor cells have the potential to be therapeutic for AD via repairing damaged endothelia in cerebrovasculature. Furthermore, we discovered that the genotypes in two genes, *KIRREL3* and *EXOC6B*, had interactive effects with the number of circulating CD34+CD133+ cells for AD risk. Interestingly, both *KIRREL3*^13, 14^ and *EXOC6* (the high homolog of *EXOC6B*)^15^ have been shown to be risk genes for and are involved in autism spectrum disorder (ASD) and AD^13–16^.

Progenitor cells have different subtypes with different surface CD expressions. Among different progenitor cells we studied, only CD34+CD133+ endothelial progenitor cells showed dose-dependent protective effects against AD development, especially in the presence of HTN and CMB (**Table 2, Figure 1**). Unlike the endothelial progenitor cells, circulating mature endothelial cells marked with CD31 expression were not associated with AD risk (**Supplement Table 1**). Supporting our findings, one clinical study (n = 40) found that AD patients had a lower level of circulating CD34+CD133+ progenitor cells than controls.^10^ Human cord blood CD133+CD34+ cells have been shown to reduce the apoptosis of endothelial and ganglion cells and increase the number of surviving CD31+ mature endothelial cells in the retina after radiation damage.^17^ CD34 (also known as sialomucin) is a transmembrane protein expressed on early hematopoietic and vascular-associated tissues.^18^ CD133 is a stem cell marker, and peripheral blood-derived CD133+ cells can further differentiate into hematopoietic, endothelial, and muscle cells *in vivo and in vitro*.^5^ Without counting CD133 expression, our study did not find a relationship between CD34+ cells and AD risk (**Supplement Table 1**), as another cross-sectional study reported that circulating CD34+ cells were higher, not lower, in AD than in controls.^19^ It is shown that the leukocyte antigen CD34 is localized to the vascular endothelium throughout the human brain,^20^ suggesting that CD34+ cells are mixed with endothelial progenitors and leukocytes, but CD34+CD133+ cells are endothelial progenitors only. On the other hand, CD133+ donor cells were detected in several vessels near areas of regeneration, where they expressed human VE-cadherin and CD31, which are biomarkers of mature endothelia.^21^ Based on our data and others, it is likely that CD34+CD133+ progenitor cells are the specific subtype of progenitors for endothelia that repair damaged brain endothelia and rescue AD risk.

CD34+CD133+ progenitor cells were not associated with different vascular diseases themselves (**Supplement Table 2**), although they were significantly associated with decreased AD risk in the presence of vascular diseases (**Table 2**). Cerebrovascular diseases are complex diseases involving multiple cell type pathologies in the brain and endothelia is one of them. The role of cerebral microvasculature pathology in AD is likely through dysfunctional endothelia (reviewed by Dalkara and Alarcon-Martinez)^22^, thus suggesting that endothelial damage/dysfunction which often occurs in cerebral pathology, but not the other cell types in cerebrovascular pathology, is a key element in AD pathogenesis. In the literature, the relationship between CD34+CD133+ cells and cerebrovascular diseases are not consistent. One study shows that autologous circulating endothelial precursor cells may have the potential for the treatment of ischemic diseases.^23^ Some studies have shown that the relationship between CD34+CD133+ progenitor cells and cerebral small vessel disease (CSVD) is positive,^24, 25^ but another one shows a negative relationship.^26^

Our study (**Table 1**) and others have shown that increased age may alter the availability of or decrease the number of circulating endothelial progenitor cells^27–29^. Some studies show that aging diseases like cardiovascular diseases^30^ and diabetes^31^ and the absence of healthy aging are associated with decreased CD34+CD133+ progenitor cells. On the other hand, an increased number of circulating CD34+CD133+ endothelial progenitor cells is associated with better outcomes of physical function^32^ and cardiovascular diseases^30^. Although we found that females have more CD34+CD133+ progenitor cells in the FHS (**Table 1**), another study found that females have a lower number of CD34+CD133+ progenitor cells than males,^33^ this discrepancy could be attributed to different age periods or some other unknown factors in the two cohorts.

By using GWAS analyses, we found that CD34+CD133+ cells differentially affected AD protection in the genotypes of two genes (*KIRREL3* and *EXOC6B*) (**Figure 2**), thus suggesting that circulating endothelial progenitor cells for AD protection work more efficiently in certain genotypes. CD34+CD133+ cells interact with *KIRREL3* SNP to reduce AD risk when the expression of *KIRREL3* was low in the brain (**Figure 5A**). One meta-analysis of GWAS with a large number of participants showed that SNPs of *KIRREL3* are a genetic risk factor for AD.^13, 14^ *KIRREL3* has been shown to mediate neuron synapse formation and cell adhesion/guidance^34–36^, as well as cell differentiation.^37, 38^ The loss of *KIRREL3* leads to changed axon organization, as well as male‒male aggression and cognitive impairment, in mice.^39–41^ Our data suggest that when brain *KIRREL3* gene expression is low and vulnerable due to its genotypes or high methylation status, the impact of circulating CD34+CD133+ progenitor cells on decreasing AD risk is increased; in the counterpart genotypes, the *KIRREL3* gene expression in the brain is high, and AD risk is not influenced by circulating endothelial CD34+CD133+ progenitor cells.

The peripheral, but not the brain, expression of *EXOC6B* was significantly and positively associated with AD risk (**Figure 5B**). In the *EXOC6B* rs61619102 CC genotype with low peripheral expression level, increased number of CD34+CD133+ progenitor cells were associated with decreased AD risk (**Figure 3** and **Table 3**). In noncarriers of *APOE* ε4, the *EXOC6* gene was found to be associated with AD risk.^15^ *EXOC6B* plays a novel role in promoting stem cell progeny differentiation^42^ and regulates cellular exocyst formation for protein-membrane trafficking and secretion.^43^ Decreased *EXOC6B* expression is associated with low insulin secretion from β cells and an increased risk of type 2 diabetes.^44^ It is plausible that the *EXOC6B* rs61619102 CC genotype regulates circulating CD34+CD133+ endothelial progenitor cells to rescue AD pathology in the brain.

Interestingly, both *KIRREL3* ^16, 35, 38, 45–50^ and *EXOC6B* ^51–53^ genes are related to ASD, another cognitive disease with a young age of onset. Like neurodegenerative diseases, ASD is also associated with cerebral hypoperfusion,^54^ suggesting the presence of common cerebrovascular pathologies in both diseases. Like AD research studies, post-mortem human studies and some animal ASD models have shown brain neuroinflammation, oxidative stress, and changes in BBB integrity.^55^ The balance between Wnt-β-catenin and Shh pathways controls angiogenesis, barrier genesis, neurodevelopment, CNS morphogenesis, and neuronal guidance that are related to both AD^56, 57^ and ASD.^55^

This study had several limitations. The FHS study was based on a non-Hispanic White population and more large cohorts with different ethnicities are needed to replicate the finding that CD34+CD133+ progenitor cells are associated with reduced risk of AD dementia. Moreover, although we had a one-time point of CD34+CD133+ cell measurement, longitudinal measurements are necessary to characterize the change of CD34+CD133+ progenitor cells during aging and the disease process. Future clinical trials with circulating CD34+CD133+ progenitor cells are key to proving that it can be therapeutic for AD prevention. Nevertheless, our study sheds light for potential cell-based therapy by using circulating CD34+CD133+ endothelial progenitors to reduce AD risk, especially among those individuals with vascular diseases and the vulnerable *KIRREL3* and *EXOC6B* genotypes.

## Data Availability

All data produced in the present study are available upon reasonable request to the authors

## Acknowledgments

This study was supported by National Institute on Aging grants U19-AG068753, RF1-AG057519, and R01-AG048927. The FHS data collection was supported by the National Heart, Lung, and Blood Institute contract (N01-HC-25195). We want to express our thanks to the FHS participants for their decades of dedication and to the FHS staff for their hard work in collecting and preparing the data.

## Author contributions

Lindsay A. Farrer and Rhoda Au guided the main mission for the U19 project and this study was part of it.

Yixuan Wang, Jinghan Huang, Ting Fang Alvin Ang, Yibo Zhu and Qiushan Tao had full access to all the data in the study and take responsibility for the integrity of the data and the accuracy of the data analysis. Yixuan Wang and Jinghan Huang conducted the main data analyses for the manuscript.

Yixuan Wang and Wei Qiao Qiu drafted the initial manuscript; Jinghan Huang and Xiaoling Zhang were actively involved in the manuscript writing.

Ting Fang Alvin Ang, Jesse Mez, Michael Alosco, Gerald V. Denis, Anna Belkina, Ashita Gurnani, Mark Ross, Bin Gong, Jingyan Han, Kathryn L. Lunetta, Thor D. Stein, Rhoda Au and Lindsay A. Farrer were critically involved in the data analyses, the manuscript writing and editing.

Xiaoling Zhang and Wei Qiao Qiu designed the study and supervised the data analyses.

## Methods

### Study samples and participants

#### Framingham Heart Study (FHS)

FHS is a single-site, multi-generation, community-based, prospective cohort study of health in Framingham, Massachusetts. The current study included 1,566 participants (*Mean_age_* = 65.93±8.91; 46.36% female) from the Offspring cohort (Gen 2) with data on genome wide single nucleotide polymorphisms (SNPs) and circulating CD34+CD133+ cells measured at examination 8 (2005-2007) who were followed subsequently for incident AD dementia. Relevant details about this cohort have been previously described.^58^ 159 of them developed incident AD through the period ending in 2019 (**Supplement Figure 1**). Informed consent was obtained from all participants and the study protocol was approved by the Institutional Review Board of Boston University Medical Campus.

#### Religious Orders Study/Memory and Aging Project (ROSMAP)

ROSMAP data were used to investigate the functions of the variants at the DNA methylation levels and gene expression levels and to explore their relationships with AD (https://dss.niagads.org/cohorts/religious-orders-study-memory-and-aging-project-rosmap/).

ROSMAP participants have annual blood draws that provide a repository of stored serum, plasma, and cells. Clinical evaluation, self-report, and medication review are used to document medical conditions. Moreover, subjects are neurologically evaluated every year, and a review of all of the ante-mortem data at the time of death leads to a final clinical diagnosis for each participant; specifically, each individual receives a diagnosis of AD, MCI, or no cognitive impairment (NCI). The details of this methodology have been described in the website.

#### Circulating endothelial progenitor cells count method

Fasting blood samples were collected from participants in order to count circulating progenitor cells and were stored at 2-8 ℃.^59^ Peripheral blood mononuclear cells (PBMCs) were used for cell phenotyping.^60^ Briefly, PBMCs were isolated via Ficoll (GE) gradient centrifugation, counted, incubated with FcR blocking antibodies (Miltenyi Biotech), labeled with anti-human KDR PE antibody (R&D Systems), anti-human CD34 FITC antibody (BD Biosciences), anti-human CD133 APC antibody (BD Biosciences). Samples were washed and fixed with 2% paraformaldehyde. Cells were analyzed on BD FACSCalibur cytometer running CellQuest software (BD Biosciences). Data were analyzed using FlowJo 8 (Flowjo, Inc) and positive populations were identified by comparisons of stained samples with samples incubated with isotype-matching IgG antibodies conjugated to FITC and PE. Compensation was performed using single-stain controls. Percentages of CD34+, CD34+CD133+, CD34+CD133-, CD34-CD133+, CD34-CD133-, CD34+, and CD34+KDR+(VEGFR2) cells were quantified in the data. Similarly, anti-human CD31 PE (BD Bioscences) and anti-human CD45 PerCP antibody (BD Biosciences) were used to analyze CD31+, CD31-, CD31+CD45-, CD31+DIM, and CD31+lymphoid cells.

#### Dementia/AD diagnoses

All of the Gen 2 participants were followed for incident AD and dementia. The Mini-Mental State Examination (MMSE) was administered beginning at the time of the 5^th^ health exam (i.e., 1991-1995) to monitor changes in cognitive status. A decrease in MMSE performance of 3 or more points from the immediately preceding exam or 5 or more points across all of the exams would indicate a change in cognitive status that warranted review by a dementia diagnostic panel consisting of at least one neurologist and one neuropsychologist. Furthermore, from 1999-2005, all of the surviving Gen 2 participants were invited for an in-depth cognitive examination, which also screened for incident cognitive impairment that warranted review by the dementia diagnostic panel. Consensus diagnostic procedures according to the criteria of Diagnostic and Statistical Manual of Mental Disorders (DSM-IV) and National Institute of Neurological and Communicative Diseases and Stroke/Alzheimer’s Disease and Related Disorders Association (NINCDS–ADRDA) for dementia and AD were conducted and have been previously described.^61^ Incidences of dementia and AD after the CD34+CD133+ measurement at Exam 8 were used for the analyses.

### Peripheral vascular diseases and cerebrovascular diseases

#### Coronary Heart Disease (CHD)

Participants were diagnosed as having developed coronary heart disease (CHD) if upon review of the case, a panel of three investigators (The Framingham Endpoint Review Committee) agreed on one of the following definite manifestations: myocardial infarction, coronary insufficiency, angina pectoris, sudden death from CHD, or nonsudden death from CHD.

#### Hypertension (HTN)

We used treatment for HTN as a proxy for having HTN. Treatment for HTN was based on evidence of prescribed hypertension medication use, self-report or FHS M.D. report of treatment for hypertension at the time of the exam. If a discrepancy between these 3 sources of information occurred, a chart review of medical records was conducted to make a final determination of HTN treatment.

#### Stroke

The diagnosis of stroke was based on the occurrence of a clinically evident stroke documented by clinical records that was reviewed by at least two neurologists. Stroke was defined as the sudden or rapid onset of a focal neurologic deficit persisting for greater than 24 hours.

#### Silent Brain Infarcts and Cerebral Microbleeds (CMB)

MRI infarcts in persons without a clinical stroke documented by the Framingham Stroke surveillance team, were considered silent brain infarct^62^ and cerebral microbleeds (CMB) were examined by visual inspection of brain MRI scans as have been previously described^63^. Exclusion criteria for the brain MRI scan included those who could not undergo MRI due to claustrophobia, presence of a pacemaker or any other source of metal in their body.

#### White Matter Hyperintensity (WMH)

White matter hyperintensity (WMH) volumetric measures were extracted from a combination of FLAIR and 3D T1 images with a modified Bayesian probability structure based on a previously published method of histogram fitting.^64^ Volumes are log-transformed to normalize population variance. Details have been described previously.^64–68^ We dichotomized WMH volume into high and low levels using the median as the cutoff.

#### Genome-wide association studies (GWAS)

To study the genotypes that were potentially impacted by circulating CD34+CD133+ progenitor cells for AD, genome-wide association studies (GWAS) were performed using the Framingham analytical pipeline. We conducted an interaction GWAS with log transformed CD34+CD133+ cell frequency by using a logistic regression model for AD. The relationship between AD and interaction terms (CD34+CD133+ frequency and genome-wide SNP dosage) was tested using GEEPACK (logistic regression utilizing generalized estimating equations) while adjusting for age, sex, years of education, and the first 10 population principal components (PCs). Additive models were assumed. Only autosomal SNPs on chromosomes 1-22 were considered. A statically filtered set of dosage files for MACH imputation via the 1,000 Genomes reference files from March 2012 of only samples of European ancestry was used. The filtering criterion was r^2^ ≥0.1. Moreover, only SNPs with minor allele frequency (MAF) ≥10% were chosen for the analysis. In addition, samples with a genotyping rate less than 97% and with an excess number of heterozygote observations or Mendelian errors were removed. Manhattan plot, QQ plot, and genomic control^69^ were used for visualization and quality control, and LocusZoom^70^ was used to present the regional information. Only p values <5.0×10^-8^ were considered to be statistically significant for the SNP-CD34+CD133+ progenitor cell interactive effects for AD.

#### Gene expression analysis and DNA methylation

We additionally explored the gene expression patterns across different AD brain regions by using Agora (https://agora.adknowledgeportal.org/genes), which included 9 brain regions, e.g. the anterior cingulate cortex (ACC), cerebellum (CBE), dorsolateral prefrontal cortex (DLPFC), frontal pole (FP), inferior frontal gyrus (IFG), posterior cingulate cortex (PCC), parahippocampal gyrus (PHG), superior temporal gyrus (STG), and temporal cortex (TCX). Two AD-related traits, e.g., BRAAK (neurofibrillary tangles) and CERAD (neuritic plaques), were also measured. In addition, GTEx Portal (https://gtexportal.org/home/) was used to study the associations between the selected SNPs and gene expression (eQTLs). The details of these procedures have been previously described.^71^

We used Brain xQTLServe from ROSMAP (http://mostafavilab.stat.ubc.ca/xQTLServe/) to explore the associations between SNPs and DNA methylation (mQTLs). Genotype data were generated from 2,093 individuals of European descent. Of these individuals, DNA methylation (450K Illumina array) (n=468) was derived from postmortem frozen samples of a single cortical region, DLPFC. Gene expression data were generated by using RNA-sequencing (Illumina HiSeq) from the DLPFC of 494 individuals at an average sequence depth of 90 M reads. The details of this methodology have been previously described.^72^

#### Statistical analysis

Data were analyzed by using R version 3.6.2. The Cox models were applied to circulating cells including CD34+CD133+, CD34+, CD34+CD133-, CD34-CD133+, CD34-CD133- or CD34+KDR+(VEGFR2), CD31+, CD31-, CD31+DIM and CD31+lymphoid cells to examine the relationship between CD34+CD133+ or quartiles and dementia/AD diagnosis risk with adjustments for age, sex, years of education, *APOE* ε4, and different vascular diseases (**Supplement Table 1**). Only CD34+CD133+ progenitor cells were found to be associated with AD dementia.

Circulating CD34+CD133+ progenitor cells were first divided into four quartiles based on cell numbers. Quartiles of cell numbers were compared on the number of subjects, age at baseline, sex, years of education, *APOE* ε4, incident AD/dementia status, CHD, HTN and different cerebrovascular diseases using analysis of variance (ANOVA), chi-squared tests and Kaplan‒Meier survival analysis. We further stratified the subjects into the absence and the presence of different vascular pathologies and examined the relationship between CD34+CD133+ quartiles and dementia/AD diagnosis risk by using the chi-squared test in each subgroup. Furthermore, the Cox model was applied to the stratification of 1) no vascular diseases, 2) peripheral vascular diseases, and 3) cerebrovascular diseases with adjustments for age, sex, years of education and *APOE* ε4. Additionally, the main effects and interaction effects (with CD34+CD133+) of GWAS-selected SNPs were obtained by using logistic regression and Cox proportional hazard regression adjusting for age, sex, years of education, *APOE* ε4, and PCs. These interaction effects were further assessed by applying stratification analyses of genotypes with Cox proportional hazard regression models and a Kaplan‒Meier survival analysis.

## Reference

1. Silva MVF, Loures CMG, Alves LCV, de Souza LC, Borges KBG, Carvalho MDG. Alzheimer’s disease: risk factors and potentially protective measures. J Biomed Sci. May 9 2019;26(1):33. doi:10.1186/s12929-019-0524-y

2. Chou RC, Kane M, Ghimire S, Gautam S, Gui J. Treatment for Rheumatoid Arthritis and Risk of Alzheimer’s Disease: A Nested Case-Control Analysis. CNS Drugs. Nov 2016;30(11):1111–1120. doi:10.1007/s40263-016-0374-z

3. Drake JD, Chambers AB, Ott BR, Daiello LA, Alzheimer’s Disease Neuroimaging I. Peripheral Markers of Vascular Endothelial Dysfunction Show Independent but Additive Relationships with Brain-Based Biomarkers in Association with Functional Impairment in Alzheimer’s Disease. J Alzheimers Dis. 2021;80(4):1553–1565. doi:10.3233/JAD-200759

4. Zhang Z, Na H, Gan Q, Tao Q, Alekseyev Y, Hu J, Yan Z, Yang JB, Tian H, Zhu S, et al. Monomeric C-reactive protein via endothelial CD31 for neurovascular inflammation in an ApoE genotype-dependent pattern: A risk factor for Alzheimer’s disease? Aging Cell. Nov 2021;20(11):e13501. doi:10.1111/acel.13501

5. Meregalli M, Farini A, Belicchi M, Torrente Y. CD133(+) cells isolated from various sources and their role in future clinical perspectives. Expert Opin Biol Ther. Nov 2010;10(11):1521–8. doi:10.1517/14712598.2010.528386

6. Rigato M, Avogaro A, Fadini GP. Levels of Circulating Progenitor Cells, Cardiovascular Outcomes and Death: A Meta-Analysis of Prospective Observational Studies. Circ Res. Jun 10 2016;118(12):1930–9. doi:10.1161/CIRCRESAHA.116.308366

7. Yamaguchi T, Kanayasu-Toyoda T, Uchida E. Angiogenic cell therapy for severe ischemic diseases. Biol Pharm Bull. 2013;36(2):176–81. doi:10.1248/bpb.b12-01008

8. Maslovaric M, Fatic N, Delevic E. State of the art of stem cell therapy for ischaemic cardiomyopathy. Part 1. Angiol Sosud Khir. 2019;25(3):39-52. Ispol’zovanie stvolovykh kletok pri ishemicheskoi kardiomiopatii. Chast’ 1. doi:10.33529/ANGIO2019324

9. Miller-Kasprzak E, Jagodzinski PP. Endothelial progenitor cells as a new agent contributing to vascular repair. Arch Immunol Ther Exp (Warsz). Jul-Aug 2007;55(4):247–59. doi:10.1007/s00005-007-0027-5

10. Kong XD, Zhang Y, Liu L, Sun N, Zhang MY, Zhang JN. Endothelial progenitor cells with Alzheimer’s disease. Chin Med J (Engl). Mar 2011;124(6):901–6.

11. Maler JM, Spitzer P, Lewczuk P, Kornhuber J, Herrmann M, Wiltfang J. Decreased circulating CD34+ stem cells in early Alzheimer’s disease: Evidence for a deficient hematopoietic brain support? Mol Psychiatry. Dec 2006;11(12):1113–5. doi:10.1038/sj.mp.4001913

12. Kutikhin AG, Sinitsky MY, Yuzhalin AE, Velikanova EA. Shear stress: An essential driver of endothelial progenitor cells. J Mol Cell Cardiol. May 2018;118:46–69. doi:10.1016/j.yjmcc.2018.03.007

13. Sun J, Song F, Wang J, Han G, Bai Z, Xie B, Feng X, Jia J, Duan Y, Lei H. Hidden risk genes with high-order intragenic epistasis in Alzheimer’s disease. J Alzheimers Dis. 2014;41(4):1039–56. doi:10.3233/JAD-140054

14. Hatcher C, Relton CL, Gaunt TR, Richardson TG. Leveraging brain cortex-derived molecular data to elucidate epigenetic and transcriptomic drivers of complex traits and disease. Transl Psychiatry. Feb 28 2019;9(1):105. doi:10.1038/s41398-019-0437-2

15. Jiang S, Zhang CY, Tang L, Zhao LX, Chen HZ, Qiu Y. Integrated Genomic Analysis Revealed Associated Genes for Alzheimer’s Disease in APOE4 Non-Carriers. Curr Alzheimer Res. 2019;16(8):753–763. doi:10.2174/1567205016666190823124724

16. Kalsner L, Twachtman-Bassett J, Tokarski K, Stanley C, Dumont-Mathieu T, Cotney J, Chamberlain S. Genetic testing including targeted gene panel in a diverse clinical population of children with autism spectrum disorder: Findings and implications. Mol Genet Genomic Med. Mar 2018;6(2):171–185. doi:10.1002/mgg3.354

17. Chen S, Li M, Sun J, Wang D, Weng C, Zeng Y, Li Y, Huo S, Huang X, Li S, et al. Human Umbilical Cord Blood-Derived CD133(+)CD34(+) Cells Protect Retinal Endothelial Cells and Ganglion Cells in X-Irradiated Rats through Angioprotective and Neurotrophic Factors. Front Cell Dev Biol. 2022;10:801302. doi:10.3389/fcell.2022.801302

18. AbuSamra DB, Aleisa FA, Al-Amoodi AS, Jalal Ahmed HM, Chin CJ, Abuelela AF, Bergam P, Sougrat R, Merzaban JS. Not just a marker: CD34 on human hematopoietic stem/progenitor cells dominates vascular selectin binding along with CD44. Blood Adv. Dec 26 2017;1(27):2799–2816. doi:10.1182/bloodadvances.2017004317

19. Bigalke B, Schreitmuller B, Sopova K, Paul A, Stransky E, Gawaz M, Stellos K, Laske C. Adipocytokines and CD34 progenitor cells in Alzheimer’s disease. PLoS One. 2011;6(5):e20286. doi:10.1371/journal.pone.0020286

20. Kalaria RN, Kroon SN. Expression of leukocyte antigen CD34 by brain capillaries in Alzheimer’s disease and neurologically normal subjects. Acta Neuropathol. 1992;84(6):606–12. doi:10.1007/BF00227737

21. Torrente Y, Belicchi M, Sampaolesi M, Pisati F, Meregalli M, D’Antona G, Tonlorenzi R, Porretti L, Gavina M, Mamchaoui K, et al. Human circulating AC133(+) stem cells restore dystrophin expression and ameliorate function in dystrophic skeletal muscle. J Clin Invest. Jul 2004;114(2):182–95. doi:10.1172/JCI20325

22. Dalkara T, Alarcon-Martinez L. Cerebral microvascular pericytes and neurogliovascular signaling in health and disease. Brain Res. Oct 14 2015;1623:3–17. doi:10.1016/j.brainres.2015.03.047

23. Untergasser G, Koeck R, Wolf D, Rumpold H, Ott H, Debbage P, Koppelstaetter C, Gunsilius E. CD34+/CD133-circulating endothelial precursor cells (CEP): characterization, senescence and in vivo application. Exp Gerontol. Jun 2006;41(6):600–8. doi:10.1016/j.exger.2006.03.019

24. Kapoor A, Gaubert A, Marshall A, Meier IB, Yew B, Ho JK, Blanken AE, Dutt S, Sible IJ, Li Y, et al. Increased Levels of Circulating Angiogenic Cells and Signaling Proteins in Older Adults With Cerebral Small Vessel Disease. Front Aging Neurosci. 2021;13:711784. doi:10.3389/fnagi.2021.711784

25. Huang ZX, Fang J, Zhou CH, Zeng J, Yang D, Liu Z. CD34(+) cells and endothelial progenitor cell subpopulations are associated with cerebral small vessel disease burden. Biomark Med. Feb 2021;15(3):191–200. doi:10.2217/bmm-2020-0350

26. Taguchi A, Matsuyama T, Moriwaki H, Hayashi T, Hayashida K, Nagatsuka K, Todo K, Mori K, Stern DM, Soma T, et al. Circulating CD34-positive cells provide an index of cerebrovascular function. Circulation. Jun 22 2004;109(24):2972–5. doi:10.1161/01.CIR.0000133311.25587.DE

27. Heiss C, Keymel S, Niesler U, Ziemann J, Kelm M, Kalka C. Impaired progenitor cell activity in age-related endothelial dysfunction. J Am Coll Cardiol. May 3 2005;45(9):1441–8. doi:10.1016/j.jacc.2004.12.074

28. Povsic TJ, Zhou J, Adams SD, Bolognesi MP, Attarian DE, Peterson ED. Aging is not associated with bone marrow-resident progenitor cell depletion. J Gerontol A Biol Sci Med Sci. Oct 2010;65(10):1042–50. doi:10.1093/gerona/glq110

29. Madonna R, Ferdinandy P, De Caterina R, Willerson JT, Marian AJ. Recent developments in cardiovascular stem cells. Circ Res. Dec 5 2014;115(12):e71–8. doi:10.1161/CIRCRESAHA.114.305567

30. Al Mheid I, Hayek SS, Ko YA, Akbik F, Li Q, Ghasemzadeh N, Martin GS, Long Q, Hammadah M, Maziar Zafari A, et al. Age and Human Regenerative Capacity Impact of Cardiovascular Risk Factors. Circ Res. Sep 16 2016;119(7):801–9. doi:10.1161/CIRCRESAHA.116.308461

31. Povsic TJ, Sloane R, Green JB, Zhou J, Pieper CF, Pearson MP, Peterson ED, Cohen HJ, Morey MC. Depletion of circulating progenitor cells precedes overt diabetes: a substudy from the VA enhanced fitness trial. J Diabetes Complications. Nov-Dec 2013;27(6):633–6. doi:10.1016/j.jdiacomp.2013.08.004

32. Povsic TJ, Sloane R, Pieper CF, Pearson MP, Peterson ED, Cohen HJ, Morey MC. Endothelial Progenitor Cell Levels Predict Future Physical Function: An Exploratory Analysis From the VA Enhanced Fitness Study. J Gerontol A Biol Sci Med Sci. Mar 2016;71(3):362–9. doi:10.1093/gerona/glv180

33. Topel ML, Hayek SS, Ko YA, Sandesara PB, Samman Tahhan A, Hesaroieh I, Mahar E, Martin GS, Waller EK, Quyyumi AA. Sex Differences in Circulating Progenitor Cells. J Am Heart Assoc. Oct 3 2017;6(10)doi:10.1161/JAHA.117.006245

34. Volker LA, Maar BA, Pulido Guevara BA, Bilkei-Gorzo A, Zimmer A, Bronneke H, Dafinger C, Bertsch S, Wagener JR, Schweizer H, et al. Neph2/Kirrel3 regulates sensory input, motor coordination, and home-cage activity in rodents. Genes Brain Behav. Nov 2018;17(8):e12516. doi:10.1111/gbb.12516

35. Taylor MR, Martin EA, Sinnen B, Trilokekar R, Ranza E, Antonarakis SE, Williams ME. Kirrel3-Mediated Synapse Formation Is Attenuated by Disease-Associated Missense Variants. J Neurosci. Jul 8 2020;40(28):5376–5388. doi:10.1523/JNEUROSCI.3058-19.2020

36. Baig DN, Yanagawa T, Tabuchi K. Distortion of the normal function of synaptic cell adhesion molecules by genetic variants as a risk for autism spectrum disorders. Brain Res Bull. Mar 2017;129:82–90. doi:10.1016/j.brainresbull.2016.10.006

37. Tamir-Livne Y, Mubariki R, Bengal E. Adhesion molecule Kirrel3/Neph2 is required for the elongated shape of myocytes during skeletal muscle differentiation. Int J Dev Biol. 2017;61(3-4-5):337–345. doi:10.1387/ijdb.170005eb

38. Martin EA, Muralidhar S, Wang Z, Cervantes DC, Basu R, Taylor MR, Hunter J, Cutforth T, Wilke SA, Ghosh A, et al. Correction: The intellectual disability gene Kirrel3 regulates target-specific mossy fiber synapse development in the hippocampus. Elife. Jun 16 2016;5doi:10.7554/eLife.18706

39. Martin EA, Woodruff D, Rawson RL, Williams ME. Examining Hippocampal Mossy Fiber Synapses by 3D Electron Microscopy in Wildtype and Kirrel3 Knockout Mice. eNeuro. May-Jun 2017;4(3)doi:10.1523/ENEURO.0088-17.2017

40. Prince JE, Brignall AC, Cutforth T, Shen K, Cloutier JF. Kirrel3 is required for the coalescence of vomeronasal sensory neuron axons into glomeruli and for male-male aggression. Development. Jun 2013;140(11):2398–408. doi:10.1242/dev.087262

41. Choi SY, Han K, Cutforth T, Chung W, Park H, Lee D, Kim R, Kim MH, Choi Y, Shen K, et al. Mice lacking the synaptic adhesion molecule Neph2/Kirrel3 display moderate hyperactivity and defective novel object preference. Front Cell Neurosci. 2015;9:283. doi:10.3389/fncel.2015.00283

42. Mao Y, Tu R, Huang Y, Mao D, Yang Z, Lau PK, Wang J, Ni J, Guo Y, Xie T. The exocyst functions in niche cells to promote germline stem cell differentiation by directly controlling EGFR membrane trafficking. Development. Jun 28 2019;146(13)doi:10.1242/dev.174615

43. TerBush DR, Maurice T, Roth D, Novick P. The Exocyst is a multiprotein complex required for exocytosis in Saccharomyces cerevisiae. EMBO J. Dec 2 1996;15(23):6483–94.

44. Sulaiman N, Yaseen Hachim M, Khalique A, Mohammed AK, Al Heialy S, Taneera J. EXOC6 (Exocyst Complex Component 6) Is Associated with the Risk of Type 2 Diabetes and Pancreatic beta-Cell Dysfunction. Biology (Basel). Mar 1 2022;11(3)doi:10.3390/biology11030388

45. Hisaoka T, Komori T, Kitamura T, Morikawa Y. Abnormal behaviours relevant to neurodevelopmental disorders in Kirrel3-knockout mice. Sci Rep. Jan 23 2018;8(1):1408. doi:10.1038/s41598-018-19844-7

46. Liu YF, Sowell SM, Luo Y, Chaubey A, Cameron RS, Kim HG, Srivastava AK. Autism and Intellectual Disability-Associated KIRREL3 Interacts with Neuronal Proteins MAP1B and MYO16 with Potential Roles in Neurodevelopment. PLoS One. 2015;10(4):e0123106. doi:10.1371/journal.pone.0123106

47. Bhalla K, Luo Y, Buchan T, Beachem MA, Guzauskas GF, Ladd S, Bratcher SJ, Schroer RJ, Balsamo J, DuPont BR, et al. Alterations in CDH15 and KIRREL3 in patients with mild to severe intellectual disability. Am J Hum Genet. Dec 2008;83(6):703–13. doi:10.1016/j.ajhg.2008.10.020

48. Ciaccio C, Leonardi E, Polli R, Murgia A, D’Arrigo S, Granocchio E, Chiapparini L, Pantaleoni C, Esposito S. A Missense De Novo Variant in the CASK-interactor KIRREL3 Gene Leading to Neurodevelopmental Disorder with Mild Cerebellar Hypoplasia. Neuropediatrics. Dec 2021;52(6):484–488. doi:10.1055/s-0041-1725964

49. Anzick S, Thurm A, Burkett S, Velez D, Cho E, Chlebowski C, Virtaneva K, Bruno D, Martin CB, Lang DM, et al. Chromoanasynthesis as a cause of Jacobsen syndrome. Am J Med Genet A. Nov 2020;182(11):2533–2539. doi:10.1002/ajmg.a.61824

50. Guerin A, Stavropoulos DJ, Diab Y, Chenier S, Christensen H, Kahr WH, Babul-Hirji R, Chitayat D. Interstitial deletion of 11q-implicating the KIRREL3 gene in the neurocognitive delay associated with Jacobsen syndrome. Am J Med Genet A. Oct 2012;158A(10):2551–6. doi:10.1002/ajmg.a.35621

51. Evers C, Maas B, Koch KA, Jauch A, Janssen JW, Sutter C, Parker MJ, Hinderhofer K, Moog U. Mosaic deletion of EXOC6B: further evidence for an important role of the exocyst complex in the pathogenesis of intellectual disability. Am J Med Genet A. Dec 2014;164A(12):3088–94. doi:10.1002/ajmg.a.36770

52. Fruhmesser A, Blake J, Haberlandt E, Baying B, Raeder B, Runz H, Spreiz A, Fauth C, Benes V, Utermann G, et al. Disruption of EXOC6B in a patient with developmental delay, epilepsy, and a de novo balanced t(2;8) translocation. Eur J Hum Genet. Oct 2013;21(10):1177–80. doi:10.1038/ejhg.2013.18

53. Borsani G, Piovani G, Zoppi N, Bertini V, Bini R, Notarangelo L, Barlati S. Cytogenetic and molecular characterization of a de-novo t(2p;7p) translocation involving TNS3 and EXOC6B genes in a boy with a complex syndromic phenotype. Eur J Med Genet. Jul-Aug 2008;51(4):292–302. doi:10.1016/j.ejmg.2008.02.006

54. Bjorklund G, Kern JK, Urbina MA, Saad K, El-Houfey AA, Geier DA, Chirumbolo S, Geier MR, Mehta JA, Aaseth J. Cerebral hypoperfusion in autism spectrum disorder. Acta Neurobiol Exp (Wars). 2018;78(1):21–29.

55. Gozal E, Jagadapillai R, Cai J, Barnes GN. Potential crosstalk between sonic hedgehog-WNT signaling and neurovascular molecules: Implications for blood-brain barrier integrity in autism spectrum disorder. J Neurochem. Oct 2021;159(1):15–28. doi:10.1111/jnc.15460

56. Liebner S, Plate KH. Differentiation of the brain vasculature: the answer came blowing by the Wnt. J Angiogenes Res. Jan 14 2010;2:1. doi:10.1186/2040-2384-2-1

57. Chen SD, Yang JL, Lin YC, Chao AC, Yang DI. Emerging Roles of Inhibitor of Differentiation-1 in Alzheimer’s Disease: Cell Cycle Reentry and Beyond. Cells. Jul 21 2020;9(7)doi:10.3390/cells9071746

58. Kannel WB, Feinleib M, McNamara PM, Garrison RJ, Castelli WP. An investigation of coronary heart disease in families. The Framingham offspring study. Am J Epidemiol. Sep 1979;110(3):281–90. doi:10.1093/oxfordjournals.aje.a112813

59. Muggeridge D, Dodd J, Ross MD. CD34(+) progenitors are predictive of mortality and are associated with physical activity in cardiovascular disease patients. Atherosclerosis. Sep 2021;333:108–115. doi:10.1016/j.atherosclerosis.2021.07.004

60. Cheng S, Wang N, Larson MG, Palmisano JN, Mitchell GF, Benjamin EJ, Vasan RS, Levy D, McCabe EL, Vita JA, et al. Circulating angiogenic cell populations, vascular function, and arterial stiffness. Atherosclerosis. Jan 2012;220(1):145–50. doi:10.1016/j.atherosclerosis.2011.10.015

61. Satizabal CL, Beiser AS, Chouraki V, Chene G, Dufouil C, Seshadri S. Incidence of Dementia over Three Decades in the Framingham Heart Study. N Engl J Med. Feb 11 2016;374(6):523–32. doi:10.1056/NEJMoa1504327

62. Das RR, Seshadri S, Beiser AS, Kelly-Hayes M, Au R, Himali JJ, Kase CS, Benjamin EJ, Polak JF, O’Donnell CJ, et al. Prevalence and correlates of silent cerebral infarcts in the Framingham offspring study. Stroke. Nov 2008;39(11):2929–35. doi:10.1161/STROKEAHA.108.516575

63. Romero JR, Beiser A, Himali JJ, Shoamanesh A, DeCarli C, Seshadri S. Cerebral microbleeds and risk of incident dementia: the Framingham Heart Study. Neurobiol Aging. Jun 2017;54:94–99. doi:10.1016/j.neurobiolaging.2017.02.018

64. DeCarli C, Miller BL, Swan GE, Reed T, Wolf PA, Garner J, Jack L, Carmelli D. Predictors of brain morphology for the men of the NHLBI twin study. Stroke. Mar 1999;30(3):529–36. doi:10.1161/01.str.30.3.529

65. Aljabar P, Heckemann RA, Hammers A, Hajnal JV, Rueckert D. Multi-atlas based segmentation of brain images: atlas selection and its effect on accuracy. Neuroimage. Jul 1 2009;46(3):726–38. doi:10.1016/j.neuroimage.2009.02.018

66. Rueckert D, Aljabar P, Heckemann RA, Hajnal JV, Hammers A. Diffeomorphic registration using B-splines. Med Image Comput Comput Assist Interv. 2006;9(Pt 2):702–9. doi:10.1007/11866763_86

67. Kochunov P, Lancaster JL, Thompson P, Woods R, Mazziotta J, Hardies J, Fox P. Regional spatial normalization: toward an optimal target. J Comput Assist Tomogr. Sep-Oct 2001;25(5):805–16. doi:10.1097/00004728-200109000-00023

68. Fletcher E, Carmichael O, Decarli C. MRI non-uniformity correction through interleaved bias estimation and B-spline deformation with a template. Annu Int Conf IEEE Eng Med Biol Soc. 2012;2012:106–9. doi:10.1109/EMBC.2012.6345882

69. Devlin B, Roeder K. Genomic control for association studies. Biometrics. Dec 1999;55(4):997–1004. doi:10.1111/j.0006-341x.1999.00997.x

70. Boughton AP, Welch RP, Flickinger M, VandeHaar P, Taliun D, Abecasis GR, Boehnke M. LocusZoom.js: Interactive and embeddable visualization of genetic association study results. Bioinformatics. Mar 17 2021;doi:10.1093/bioinformatics/btab186

71. Consortium GT. The Genotype-Tissue Expression (GTEx) project. Nat Genet. Jun 2013;45(6):580–5. doi:10.1038/ng.2653

72. Ng B, White CC, Klein HU, Sieberts SK, McCabe C, Patrick E, Xu J, Yu L, Gaiteri C, Bennett DA, et al. An xQTL map integrates the genetic architecture of the human brain’s transcriptome and epigenome. Nat Neurosci. Oct 2017;20(10):1418–1426. doi:10.1038/nn.4632

