## Supplemental Table 1-6, Supplemental Figure 1-3 for "Circulating Endothelial Progenitor Cells Reduce the Risk of Alzheimer’s Disease"

**Supplemental information**

**Supplement Table 1. Examination of the relationship between different progenitor or mature endothelial cells (log transformed) and the risk of AD/dementia by using Cox proportional hazards regression models after adjusting for age, sex, years of education, and *APOE* ε4**

| **Circulating cells** | **Alzheimer’s disease** | | **Dementia** | |
| --- | --- | --- | --- | --- |
|  | **HR (95% CI)** | **P value** | **HR (95% CI)** | **P value** |
| **CD34+CD133+** | 0.67 (0.46, 0.96)  n = 1566 | **0.03** | 0.65 (0.48, 0.90)  n = 1589 | **0.008** |
| **CD34+CD133-** | 0.73 (0.46, 1.14)  n = 1566 | 0.17 | 0.70 (0.47, 1.03)  n = 1589 | 0.07 |
| **CD34-CD133+** | 0.55 (0.28, 1.08)  n = 1566 | 0.08 | 0.80 (0.46, 1.39)  n = 1589 | 0.42 |
| **CD34-CD133-** | 382.92 (0.99, 1.48e+05)  n = 1566 | 0.05 | 65.66 (0.50, 8.63e+03)  n = 1589 | 0.09 |
| **CD34+** | 0.66 (0.40, 1.10)  n = 1667 | 0.11 | 0.66 (0.43, 1.01)  n = 1689 | 0.06 |
| **CD34+/KDR+** | 0.93 (0.66, 1.29)  n = 1668 | 0.65 | 0.89 (0.67, 1.19)  n = 1690 | 0.44 |
| **CD31+/CD45-** | 0.84 (0.62, 1.15)  n = 1699 | 0.29 | 0.81 (0.62, 1.06)  n = 1722 | 0.13 |
| **CD31+** | 0.86 (0.44, 1.67)  n = 1657 | 0.66 | 0.72 (0.42, 1.26)  n = 1679 | 0.25 |
| **CD31-** | 1.41 (0.68, 2.91)  n = 1657 | 0.35 | 1.64 (0.86, 3.12)  n = 1679 | 0.13 |
| **CD31+DIM** | 0.82 (0.37, 1.80)  n = 1657 | 0.62 | 0.78 (0.40, 1.54)  n = 1679 | 0.48 |
| **CD31+Lymphoid** | 2.06 (0.18, 23.22)  n = 1657 | 0.56 | 3.20 (0.38, 26.87)  n = 1679 | 0.28 |

**Supplement Table 2. Examination of the relationship between CD34+CD133+ (log transformed) and the risk of AD/dementia risk by using Cox proportional hazards regression models adjusted for covariates including different vascular diseases**

| **Covariates** | **Alzheimer’s disease** | |  | **Dementia** | |
| --- | --- | --- | --- | --- | --- |
|  | **HR (95% CI)** | **P value** |  | **HR (95% CI)** | **P value** |
| **Model 1: no covariates** | 0.57 (0.40, 0.81) | 0.002 |  | 0.57 (0.42, 0.77) | 2.8e-04 |
| **Model 2: Age, sex, years of education** | 0.64 (0.45, 0.93) | 0.02 |  | 0.64 (0.47, 0.87) | 0.005 |
| **Model 3: Model 2 + *APOE* ε4** | 0.67 (0.46, 0.96) | 0.03 |  | 0.65 (0.48, 0.90) | 0.008 |
| **Model 3 plus** |  | | | | |
| **CHD** | 0.67 (0.46, 0.96) | 0.03 |  | 0.65 (0.48, 0.90) | 0.008 |
| **HTN** | 0.65 (0.45, 0.94) | 0.02 |  | 0.64 (0.47, 0.88) | 0.006 |
| **Stroke** | 0.68 (0.47, 0.99) | 0.04 |  | 0.67 (0.49, 0.92) | 0.01 |
| **ABI** | 0.71 (0.48, 1.04) | 0.08 |  | 0.69 (0.50, 0.95) | 0.02 |
| **WMHI** | 0.58 (0.34, 0.99) | 0.05 |  | 0.58 (0.37, 0.90) | 0.01 |
| **CBI, large** | 0.54 (0.35, 0.85) | 0.007 |  | 0.52 (0.35, 0.76) | 6.6e-04 |
| **CBI, small** | 0.55 (0.35, 0.85) | 0.008 |  | 0.52 (0.36, 0.77) | 8.3e-04 |
| **CBI, basal ganglia** | 0.63 (0.40, 0.99) | 0.05 |  | 0.60 (0.41, 0.89) | 0.01 |
| **CMB** | 0.63 (0.38, 1.05) | 0.08 |  | 0.59 (0.39, 0.90) | 0.01 |
| **Deep CMB** | 0.61 (0.37, 1.03) | 0.06 |  | 0.59 (0.39, 0.90) | 0.02 |
| **Frontal Cortex CMB** | 0.61 (0.34, 1.10) | 0.10 |  | 0.58 (0.36, 0.94) | 0.03 |

Abbreviations: HR, hazard ratio; CHD, coronary heart disease; HTN, hypertension; ABI, atherothrombotic infarction; FLAIR WMHI, total white matter hyperintensity segmented from FLAIR sequence; CBI, covert brain infarct; CMB, cerebral microbleed.

**Supplemental Table 3. Other SNPs from GWAS interaction results of *KIRREL3* and *EXOC6B***

| **Gene/Closest gene** | **Chr** | **Position ((GRCh37)** | **Major Allele** | **Minor Allele** | **rs ID** | **Beta** | **se** | **P value** |
| --- | --- | --- | --- | --- | --- | --- | --- | --- |
| ***KIRREL3*** | | | | | | | | |
| *KIRREL3* | 11 | 126436966 | C | T | rs605226 | 1.6199 | 0.2677 | 1.60e-08 |
| *KIRREL3* | 11 | 126436922 | G | T | rs605162 | 1.6169 | 0.2678 | 1.70e-08 |
| *KIRREL3* | 11 | 126437834 | T | A | rs578463 | 1.4686 | 0.2458 | 2.40e-08 |
| *KIRREL3* | 11 | 126437941 | G | A | rs619996 | 1.4646 | 0.2452 | 2.40e-08 |
| *KIRREL3* | 11 | 126439019 | C | T | rs634964 | 1.7946 | 0.3006 | 2.50e-08 |
| ***EXOC6B*** | | | | | | | | |
| *EXOC6B* | 2 | 73082263 | G | A | rs55802296 | 1.4881 | 0.2686 | 2.30e-07 |
| *EXOC6B* | 2 | 73079675 | T | C | rs12614041 | 1.4343 | 0.2594 | 2.44e-07 |
| *EXOC6B* | 2 | 73079965 | T | A | rs56711961 | 1.4348 | 0.2596 | 2.45e-07 |
| *EXOC6B* | 2 | 73079989 | G | C | rs56181835 | 1.4349 | 0.2596 | 2.45e-07 |
| *EXOC6B* | 2 | 73081791 | T | A | rs12615875 | 1.4381 | 0.2602 | 2.45e-07 |
| *EXOC6B* | 2 | 73080639 | G | C | rs2063168 | 1.4556 | 0.2643 | 2.70e-07 |
| *EXOC6B* | 2 | 73076311 | T | C | rs4852897 | 1.4127 | 0.2570 | 2.84e-07 |
| *EXOC6B* | 2 | 73075651 | T | C | rs57264301 | 1.4119 | 0.2569 | 2.86e-07 |
| *EXOC6B* | 2 | 73075644 | A | G | rs59615613 | 1.4114 | 0.2568 | 2.86e-07 |
| *EXOC6B* | 2 | 73075206 | C | T | rs67919884 | 1.4057 | 0.2559 | 2.90e-07 |
| *EXOC6B* | 2 | 73069299 | C | T | rs12619068 | 1.3876 | 0.2530 | 3.04e-07 |
| *EXOC6B* | 2 | 73069973 | G | T | rs1876488 | 1.3875 | 0.2530 | 3.04e-07 |
| *EXOC6B* | 2 | 73068205 | T | A | rs67927720 | 1.3876 | 0.2530 | 3.04e-07 |
| *EXOC6B* | 2 | 73065894 | C | T | rs2135983 | 1.3875 | 0.2530 | 3.04e-07 |
| *EXOC6B* | 2 | 73073494 | A | C | rs4852896 | 1.3855 | 0.2527 | 3.04e-07 |
| *EXOC6B* | 2 | 73072728 | C | T | rs60584321 | 1.3855 | 0.2527 | 3.04e-07 |
| *EXOC6B* | 2 | 73071304 | A | T | rs12614455 | 1.3896 | 0.2534 | 3.04e-07 |
| *EXOC6B* | 2 | 73062631 | G | C | rs11126386 | 1.3878 | 0.2531 | 3.05e-07 |
| *EXOC6B* | 2 | 73064491 | G | A | rs6707107 | 1.3878 | 0.2531 | 3.05e-07 |
| *EXOC6B* | 2 | 73061305 | C | T | rs58448780 | 1.3876 | 0.2531 | 3.07e-07 |
| *EXOC6B* | 2 | 73061116 | G | A | rs4564798 | 1.3876 | 0.2531 | 3.07e-07 |
| *EXOC6B* | 2 | 73060352 | T | G | rs7577528 | 1.3876 | 0.2532 | 3.11e-07 |
| *EXOC6B* | 2 | 73060015 | A | G | rs7586886 | 1.3875 | 0.2532 | 3.11e-07 |
| *EXOC6B* | 2 | 72998061 | T | C | rs6716335 | 1.4199 | 0.2593 | 3.15e-07 |
| *EXOC6B* | 2 | 72998075 | C | T | rs6761253 | 1.4194 | 0.2592 | 3.15e-07 |
| *EXOC6B* | 2 | 73034553 | C | T | rs11126382 | 1.3861 | 0.2537 | 3.34e-07 |
| *EXOC6B* | 2 | 73072819 | A | G | rs112345272 | 1.4133 | 0.2588 | 3.39e-07 |
| *EXOC6B* | 2 | 73030396 | A | G | rs10204141 | 1.3839 | 0.2534 | 3.41e-07 |
| *EXOC6B* | 2 | 73030674 | C | T | rs10496187 | 1.3840 | 0.2535 | 3.42e-07 |
| *EXOC6B* | 2 | 73027972 | A | T | rs12619565 | 1.3835 | 0.2534 | 3.43e-07 |
| *EXOC6B* | 2 | 73013879 | T | A | rs7607226 | 1.3819 | 0.2532 | 3.45e-07 |
| *EXOC6B* | 2 | 73013836 | C | T | rs7593084 | 1.3819 | 0.2532 | 3.45e-07 |
| *EXOC6B* | 2 | 73006888 | A | G | rs4597553 | 1.3897 | 0.2547 | 3.49e-07 |
| *EXOC6B* | 2 | 73003486 | C | T | rs58890506 | 1.3998 | 0.2566 | 3.49e-07 |
| *EXOC6B* | 2 | 72997442 | R | D |  | 1.4036 | 0.2573 | 3.49e-07 |
| *EXOC6B* | 2 | 73005579 | T | C | rs970577 | 1.3899 | 0.2548 | 3.50e-07 |
| *EXOC6B* | 2 | 73007522 | T | C | rs2068410 | 1.3889 | 0.2546 | 3.50e-07 |
| *EXOC6B* | 2 | 73029379 | A | G | rs7586339 | 1.3936 | 0.2564 | 3.88e-07 |
| *EXOC6B* | 2 | 73082129 | C | T | rs11886503 | 1.4540 | 0.2680 | 4.07e-07 |
| *EXOC6B* | 2 | 73082562 | G | T | rs4852899 | 1.4756 | 0.2731 | 4.51e-07 |
| *EXOC6B* | 2 | 73082563 | C | A | rs4852900 | 1.4760 | 0.2732 | 4.51e-07 |
| *EXOC6B* | 2 | 73023457 | T | C | rs111636941 | 1.3785 | 0.2551 | 4.52e-07 |
| *EXOC6B* | 2 | 73071303 | R | D |  | 1.4171 | 0.2635 | 5.11e-07 |
| *EXOC6B* | 2 | 73082956 | T | A | rs7567893 | 1.4785 | 0.2785 | 7.12e-07 |

**Supplemental Table 4. Stratified genotype analysis of CD34+CD133+ (log transformed) on AD incidences by using Cox proportional hazards regression model adjusted by age, sex, and years of education, as well as *APOE* ε4 and PCs in FHS**

| **SNP (Gene)** | **Genotype** | **HR (95% CI)** | **P value** |
| --- | --- | --- | --- |
| rs4144611 (***KIRREL3)*** | TT | 0.29 (0.15-0.57) | **4.0e-04** |
|  | GG+TG | 1.13 (0.68-1.89) | 0.64 |
| rs580382 (***KIRREL3)*** | CC | 0.31 (0.17-0.57) | **2.0e-04** |
|  | TT+CT | 1.55 (0.89-2.68) | 0.12 |
| rs61619102 (***EXOC6B)*** | CC | 0.49 (0.31-0.75) | **1.2e-03** |
|  | GG+GC | 2.72 (1.17-6.28) | 0.02 |

**Supplement Table 5. Circulating CD34+CD133+ cells for Alzheimer’s disease in the context of genetic background in FHS, stratified by each genotype (additive) adjusting for age, sex, years of education, *APOE* ɛ4, PCs**

| **rs4144611 (*KIRREL3*)** | | | | **rs580382 (*KIRREL3*)** | | | | **rs61619102 (*EXOC6B*)** | | | |
| --- | --- | --- | --- | --- | --- | --- | --- | --- | --- | --- | --- |
| **genotype** | **CD34+CD133+ cutoffs** | **HR**  **(95% CI)** | **P value** | **genotype** | **CD34+CD133+ cutoffs** | **HR**  **(95% CI)** | **P value** | **genotype** | **CD34+CD133+ cutoffs** | **HR**  **(95% CI)** | **P value** |
| **TT**  (N=532,  AD=24) | 25% | 0.18  (0.07-0.46) | **3.3e-04** | **CC**  (N=598,  AD=29) | 25% | 0.21  (0.09-0.47) | **1.7e-04** | **CC**  (N=1023,  AD=44) | 25% | 0.48  (0.26-0.88) | **0.02** |
|  | 50% | 0.11  (0.03-0.42) | **0.001** |  | 50% | 0.16  (0.05-0.47) | **9.6e-04** |  | 50% | 0.38  (0.19-0.74) | **0.005** |
|  | 75% | *N/A*  (Zero AD) | **0.006*** |  | 75% | *N/A*  (Zero AD) | **0.002^a^** |  | 75% | 0.25  (0.08-0.81) | **0.02** |
| **TG**  (N=702,  AD=22) | 25% | 0.71  (0.29-1.75) | 0.46 | **CT**  (N=670,  AD=17) | 25% | 1.35  (0.43-4.23) | 0.60 | **GC**  (N=372,  AD=10) | 25% | 1.50  (0.27-8.45) | 0.64 |
|  | 50% | 0.80  (0.34-1.88) | 0.60 |  | 50% | 1.23  (0.46-3.27) | 0.68 |  | 50% | 7.92  (1.39-45.20) | **0.02** |
|  | 75% | 1.16  (0.43-3.10) | 0.77 |  | 75% | 1.65  (0.60-4.58) | 0.33 |  | 75% | 5.31  (1.39-20.28) | **0.01** |
| **GG**  (N=208,  AD=10) | 25% | 4.17  (0.39-44.56) | 0.24 | **TT**  (N=174,  AD=10) | 25% | 3.82  (0.35-41.05) | 0.27 | **GG^b^**  (N=47,  AD=2) | 25% | 0.00  (0.00-lnf) |  |
|  | 50% | 20.27  (1.71-240.88) | **0.02** |  | 50% | 25.55  (2.26-288.65) | **0.009** |  | 50% | Infinite  (0.00-lnf) |  |
|  | 75% | 2.31  (0.53-10.13) | 0.27 |  | 75% | 4.05  (0.84-19.55) | 0.08 |  | 75% | 0.39  (0.00-lnf) |  |

Cox proportional hazards regression model, AD incidence ~ (CD34+CD133+ cutoffs) + age + sex + years of education + *APOE* ɛ4 + PCs, stratified by genotype.

a. When CD34+CD133+ higher than 75% percentile, there are no AD cases among these genotypes, so log-rank statistics is used instead to compare groups.

b. Not enough subjects to perform the analysis.

**Supplement Table 6. Circulating CD34+CD133+ cells for Alzheimer’s disease in the context of genetic background in FHS stratified by genotypes**

|  |  |  | **Blood CD34+CD133+ Endothelia Progenitor Cells frequency, %** | | | | **P-value^a^** |
| --- | --- | --- | --- | --- | --- | --- | --- |
| **SNP (Gene)** | **Genotype** | **Overall**  **[0.002,0.609]** | **1st Q**  **[0.002,0.020]** | **2nd Q**  **[0.020,0.032]** | **3rd Q**  **[0.032,0.049]** | **4th Q**  **[0.049,0.609]** |  |
|  |  | AD, n (%) | AD, n (%) | AD, n (%) | AD, n (%) | AD, n (%) |  |
| **rs4144611 (*KIRREL3*)** | TT | 35 (6.23%) | 20 (13.25%) | 8 (5.52%) | 7 (4.90%) | 0 (0.00%) | **8.8e-05** |
|  | TG | 30 (4.11%) | 13 (6.95%) | 4 (2.50%) | 6 (3.02%) | 7 (3.83%) | 0.14 |
|  | GG | 14 (6.45%) | 1 (1.64%) | 4 (7.41%) | 5 (10.00%) | 4 (7.69%) | 0.30 |
| **rs580382 (*KIRREL3*)** | CC | 40 (6.36%) | 24 (14.55%) | 8 (5.10%) | 8 (4.85%) | 0 (0.00%) | **2.1e-06** |
|  | CT | 28 (4.01%) | 9 (4.86%) | 5 (3.21%) | 6 (3.24%) | 8 (4.62%) | 0.79 |
|  | TT | 11 (6.11%) | 1 (2.04%) | 3 (6.52%) | 4 (9.52%) | 3 (6.98%) | 0.51 |
| **rs61619102 (*EXOC6B*)** | CC | 59 (5.53%) | 31 (10.23%) | 13 (5.51%) | 10 (3.70%) | 5 (1.94%) | **1.1e-04** |
|  | GC | 18 (4.60%) | 3 (3.57%) | 3 (2.68%) | 7 (6.48%) | 5 (5.75%) | 0.52 |
|  | GG | 2 (4.08%) | 0 (0.00%) | 0 (0.00%) | 1 (7.14%) | 1 (8.33%) | 0.60 |

a. Chi-squared test p-value.

**Supplement Figure 1. Flow chart**

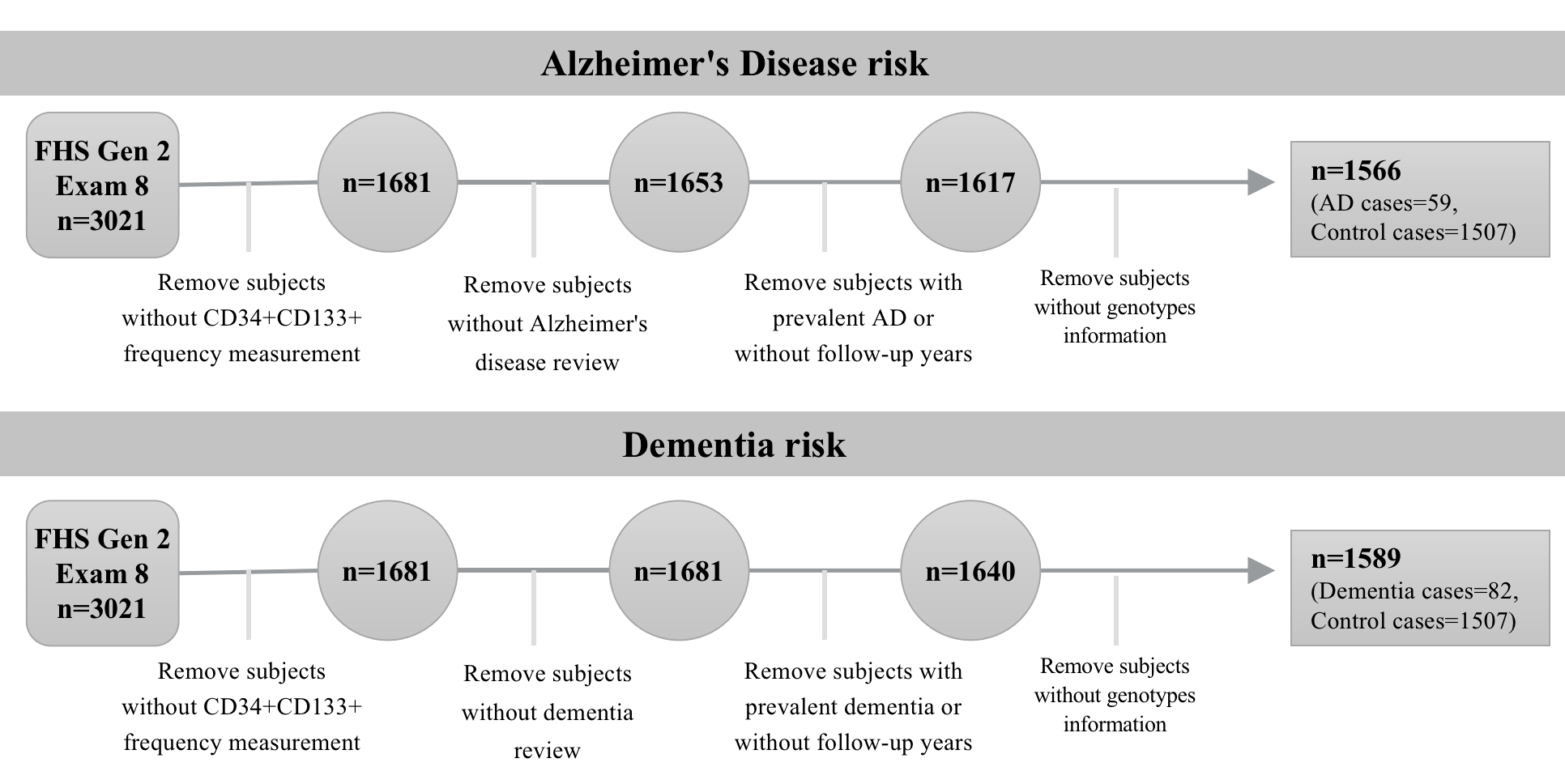

**Supplement Figure 2. Q-Q plot of the GWAS analyses**

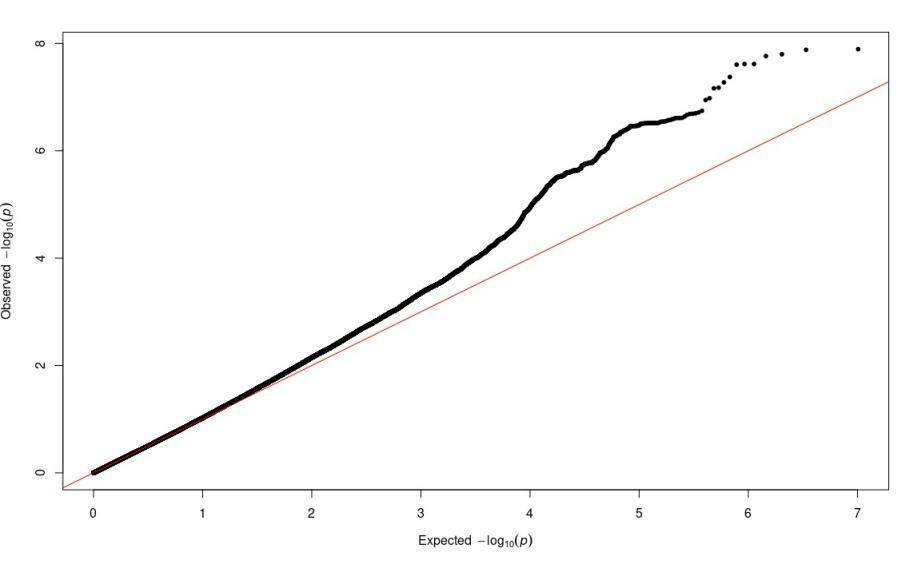

Genomic inflation (λ=1.15 before genomic control)

**Supplement Figure 3. The relationship between *KIRREL3* expression, SNPs and methylation in the ROSMAP study**

A. **The relationship between *KIRREL3* expression and the methylation site cg11751545 in the DLPFC region**

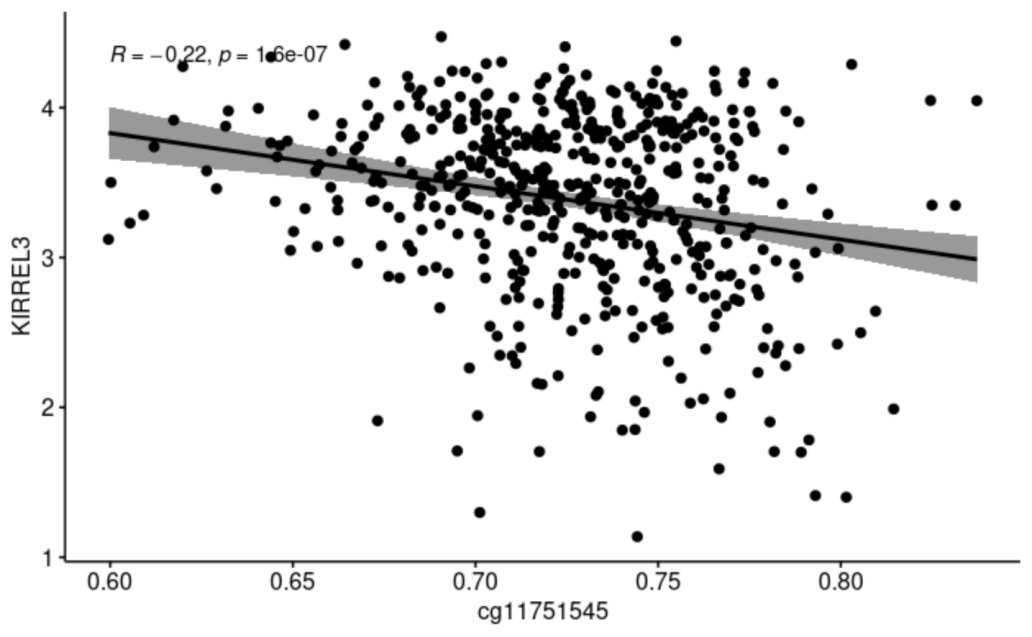

B. **The relationships between SNPs and DLPFC methylation in ROSMAP**

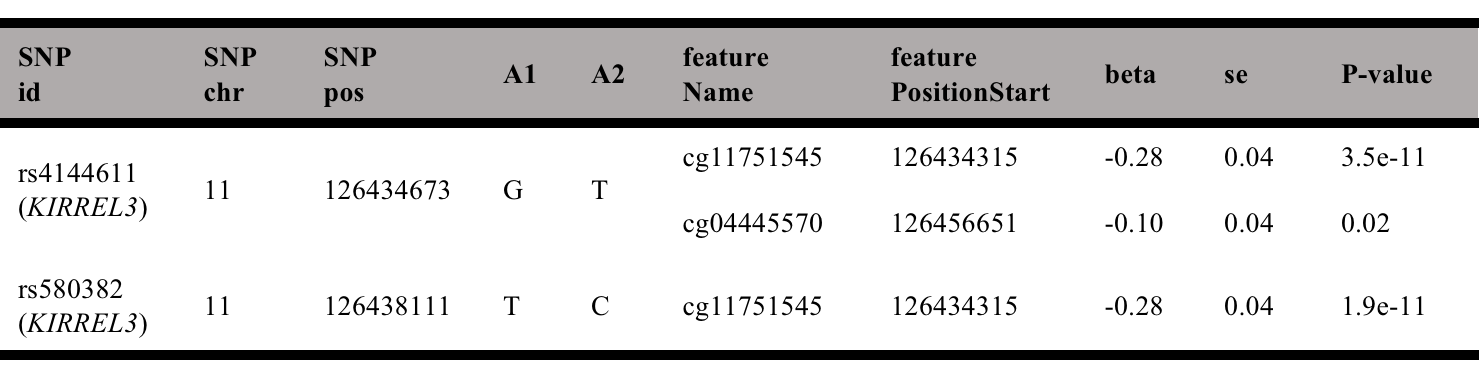
